# Medial temporal lobe hyperconnectivity is key to Alzheimer’s disease: Insight from physiological aging to dementia

**DOI:** 10.1101/2023.11.23.23298948

**Authors:** Léa Chauveau, Brigitte Landeau, Sophie Dautricourt, Anne-Laure Turpin, Marion Delarue, Oriane Hébert, Vincent de La Sayette, Gaël Chételat, Robin de Flores

## Abstract

Curing Alzheimer’s disease (AD) remains hampered by an incomplete understanding of its pathophysiology and progression. Dysfunction within medial temporal lobe networks may provide key insights, as AD proteins seem to propagate specifically through the anterior-temporal (AT) and posterior-medial (PM) systems. Using monocentric longitudinal data from 267 participants spanning physiological aging to the full AD continuum, we found that advancing age was associated with decreased PM connectivity and increased AT connectivity over adult life. When specifically assessing AD-relevant connectivity changes, all AD-associated clinicopathological features, including elevated amyloid burden, AD-typical glucose hypometabolism, hippocampal atrophy, greater cognitive impairment and faster progression from MCI to AD-dementia, were consistently linked to AT hyperconnectivity in healthy to AD-demented older adults. Our comprehensive approach allowed us to reveal that excessive connectivity within the AT network is a pivotal mechanism catalysing pathological process and progression of AD. Such findings hold promise for early diagnosis and therapeutic strategies targeting these specific network alterations.

## Introduction

Although Alzheimer’s disease (AD) is the leading cause of dementia worldwide, its biological mechanisms remain unclear, and improving its diagnosis and prognosis is crucially needed for patient care.^1^ To address these challenges, numerous studies have focused on medial temporal lobe regions, as they are the main target of neurofibrillary tangles and neuronal loss in typical AD.^2–5^ This intensive inquiry also arose from their involvement in a wide range of cognitive functions, such as declarative memory, affective processing, or spatial navigation,^6–9^ and led to the extensive use of medial temporal atrophy as a primary imaging biomarker for supporting AD diagnosis.^10,11^ However, this isolated measure lacks sensitivity for detecting preclinical changes^4^ and specificity, as many other conditions involve hippocampal shrinkage.^12–14^

Previous research has described network dysfunctions in AD that may precede structural and molecular damage, prior to symptom onset.^15–17^ Studies also showed that experimental or behavioural interventions targeting network abnormalities could enhance cognitive abilities in patients at risk and mouse models.^18^ More recently, connectome-based evidences supported prior histological and animal findings arguing that disease progression is related to network organization.^2,16,18–20^ Networks are not solely depicted as passive conduits through which pathology propagates; they may also act as independent drivers that actively modulate the spread of pathological proteins.^21,22^

Interestingly, two functional networks have been identified at the crossroad of the medial temporal lobe: the anterior-temporal (AT) and posterior-medial (PM) systems.^6^ These networks appear particularly important for AD pathophysiology due to their specific exposure to AD lesions, with tau pathology specifically targeting AT and amyloid plaques targeting PM.^23,24^ Prior research on AD has already reported abnormalities in these networks, however, results are inconsistent due to disparate methodologies and failure to consider individual differences and long-term outcomes. While reduced functional connectivity has been quite consistently described for PM, results for AT are less clear, with either increased or reduced functional connectivity in AD patients.^25–29^ Connectivity changes within the AT and PM networks have also been linked to CSF p-tau levels, white matter intensity, entorhinal thickness and memory performance in early AD.^26^ Although these AD-related markers were mostly associated with reduced AT or PM connectivity, the specific regions affected and within/between-network dynamics were unclear, which poses challenges for clinical applications. To date, there has been no comprehensive examination of functional changes in AD-sensitive networks and their relationships with markers of AD-associated brain damage and clinical advancement across the entire AD continuum, based on longitudinal data to accurately forecast disease progression. Additionally, age-specific effects have been largely underevaluated and rarely balanced against pathological mechanisms.^30–33^ These inadequacies make it challenging to identify a reliable AD-signature network dysfunction.

As functional alterations in the AT and PM networks may prompt the spread of pathology and cognitive impairment related to AD, the specific changes in each network and their clinical relevance across AD stages need to be elucidated and reconciled with age-specific effects. Here, we leveraged longitudinal monocentric data, combining neuroimaging and cognitive assessments in individuals ranging from young to older adults without cognitive impairment to the full AD continuum. The study presents a comprehensive overview of functional changes within the AT and PM networks across the whole sample, and their associations with multiple AD-relevant hallmarks and outcomes. Our findings allow to clearly identify AD-signature connectivity changes, showing that all AD-related biomarkers associated with AD pathology or clinical staging were consistently associated with AT hyperconnectivity.

## Results

### Participants

The study included 267 participants from the IMAP+ trial (https://clinicaltrials.gov/ct2/show/NCT01638949) with up to 4 years of exam follow-up. Two hundred and fifteen were cognitively unimpaired adults aged from 19 to 85 years, including 56 participants aged over 60 years without amyloid-β (Aβ) pathology, and 52 were patients with amnestic mild cognitive impairment (MCI, n = 26) or AD-dementia (n = 26) showing evidence for Aβ pathology based on PET. In the MCI group, 69% (18/26) progressed to AD-dementia within 7 years -comprising post-study follow-up. Sample characteristics are reported in Table 1.

**Table 1.**
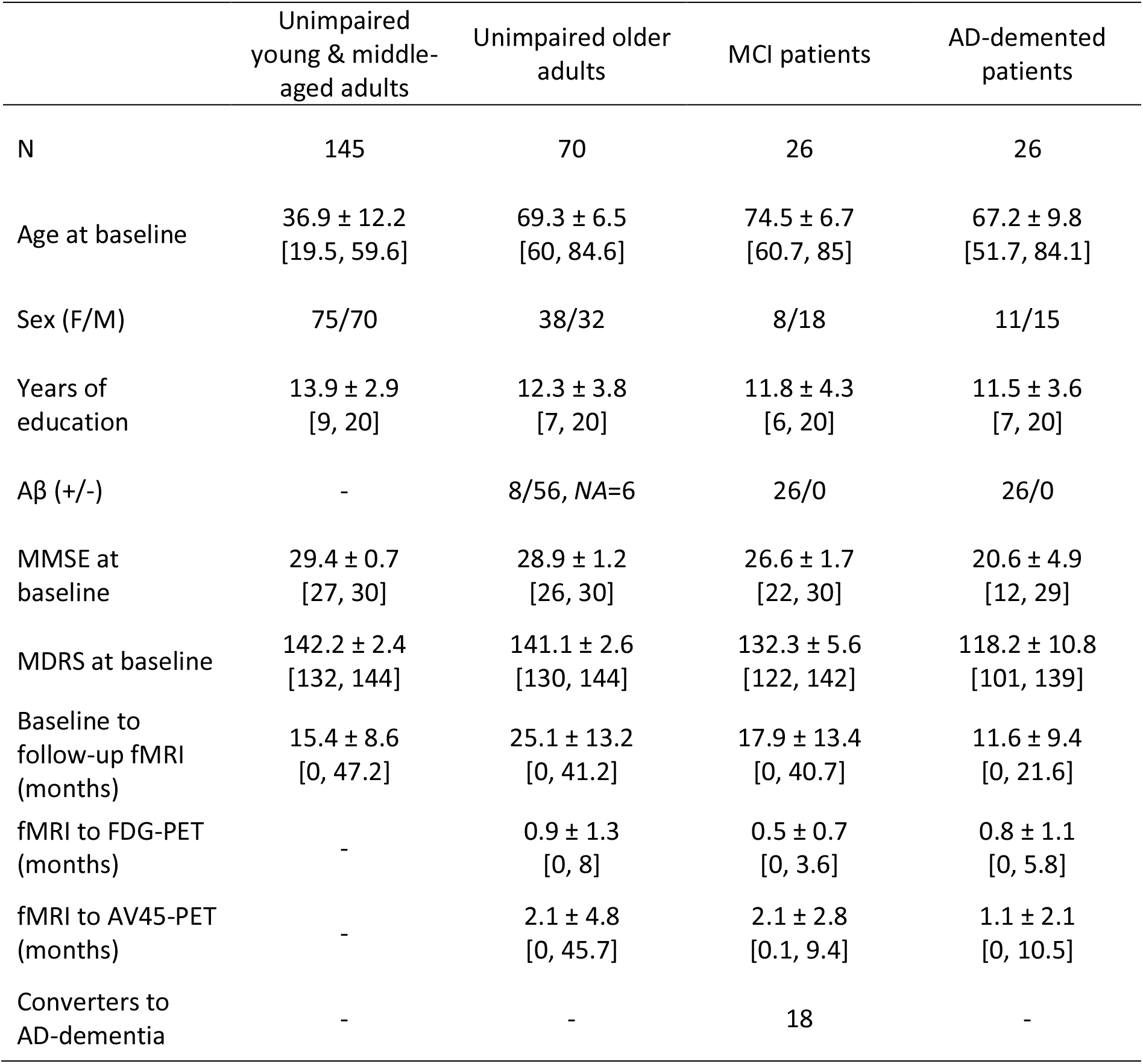
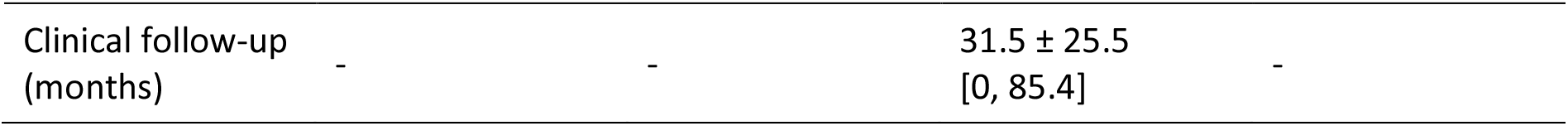
Participant characteristics. Continuous variables are indicated as follows: mean ± SD [min, max], AD: Alzheimer’s disease, MCI: mild cognitive impairment, MDRS: Mattis dementia rating scale, MMSE: Mini-mental state examination.

### Identification of the AT and PM networks

The AT and PM networks were defined following the procedure described in Fig. 1 and <u>Methods</u>. Connectivity maps were generated from seed-to-voxels correlation analyses applied to resting-state fMRI scans. As they were best suited to accurately discriminate the networks,^6^ the perirhinal cortex was used as a seed for AT and the parahippocampal cortex for PM, resulting in 2 connectivity maps per scan. Individual seeds were derived from ASHS-T1 segmentations to account for dura mater confounding and inter-individual anatomical variability in the medial temporal lobe.^34^ In order to determine AT- and PM-specific regions, perirhinal and parahippocampal functional connectivity maps were compared with paired permutation t-tests using the FSL randomise command with threshold-free cluster enhancement (TFCE) and family-wise error (FWE) correction.^35^ Output statistical maps were thresholded at 95% to create a mask for each network -that could be used in further analyses to restrict the investigation to the network of interest. These masks were obtained from the cognitively unimpaired populations relevant to the analysis, to reflect networks in physiological conditions. As such, masks intended for age-related analyses were obtained from baseline data of cognitively unimpaired adults under the age of 40, whereas masks intended for AD-related analyses were obtained from baseline data of Aβ-negative cognitively unimpaired adults over the age of 60. The corresponding masks are illustrated in Fig. 1. For both masks, the identified networks intersected the hippocampus along its longitudinal axis and consisted of previously described associated regions, including the amygdala, and the lateral entorhinal, temporopolar and orbitofrontal cortices for AT, and the retrosplenial, posterior cingulate, precuneus, medial entorhinal and ventromedial prefrontal cortices, and the angular, fusiform and inferior gyri for PM.^6,36^ Interestingly, younger adults’ masks were more extended than those of older adults, implying a loss of spatially connected regions with age (32566 versus 23343 voxels included for AT [dice = 0.71] and 67120 versus 63167 for PM [dice = 0.78]). Additional regions were located in the parietal lobules, frontal gyri and basal ganglia for AT, and in the occipital, superior frontal and postcentral gyri for PM. The average functional connectivity within each mask was then extracted for all available scans to reflect the overall functional connectivity of the AT and PM networks.

**Figure 1.**
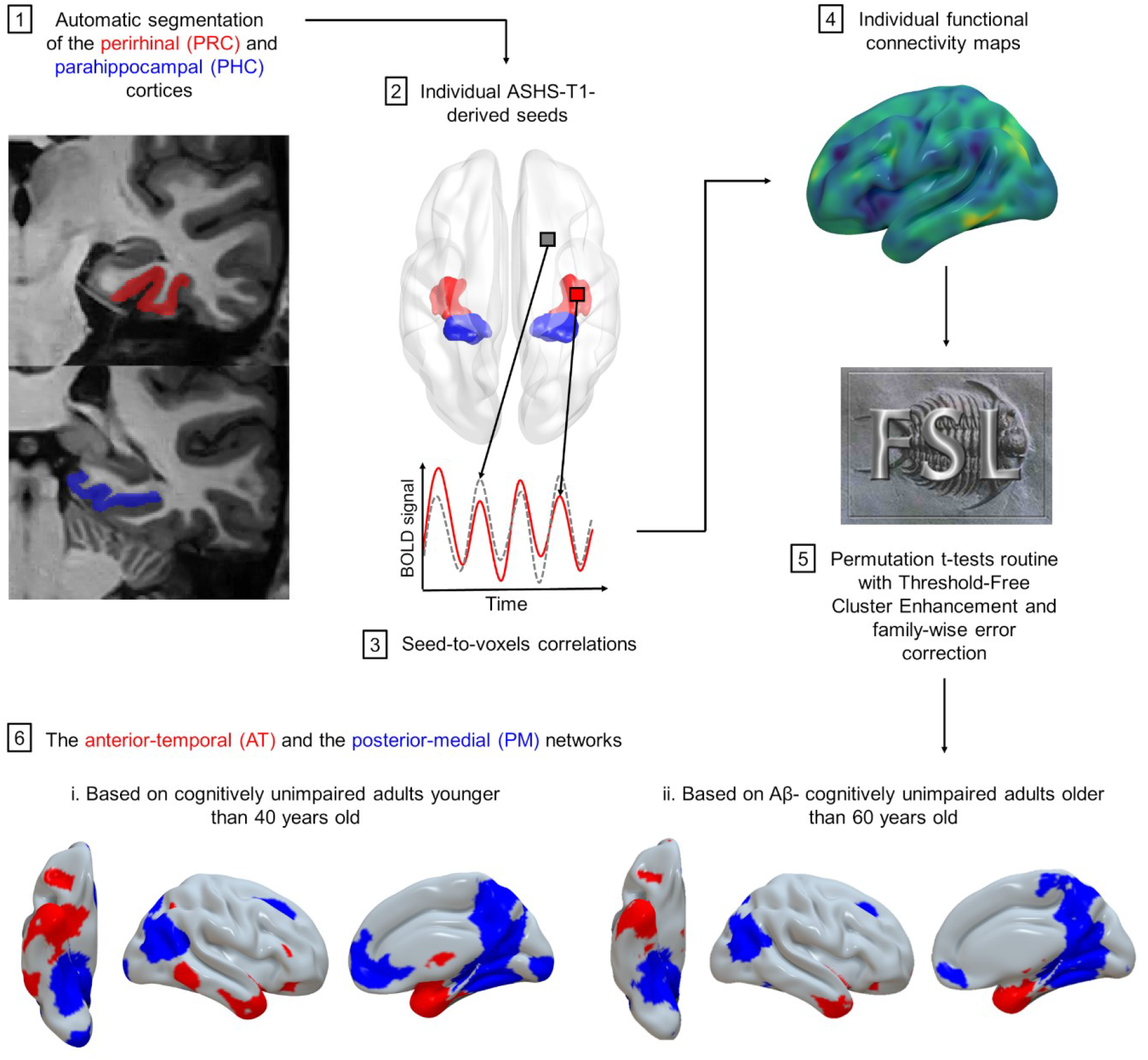
The Anterior-temporal (AT) and posterior-medial (PM) networks. Summary of the identification procedure steps described in Methods.

### Age-related changes over the adult lifespan

Age trajectories for AT and PM functional changes throughout adult life were first investigated among all cognitively unimpaired individuals using generalized additive mixed models, which provided no support for nonlinearity (edf = 1). Linear mixed models adjusted for sex and education indicated that AT functional connectivity increased with age (*F* = 5.8, *p* < 0.05, 95% CI = 5.9e^−5^, 6.1e^−4^), while PM functional connectivity decreased (*F* = 29.2, *p* < 0.001, 95% CI = -1.0e^−3^, -4.7e^−4^) (see Fig. 2, n = 214, data = 417). Additional exploratory analyses were conducted to investigate the effects of age more locally. Masks were split into regions of interest (ROIs) using the Brainnetome atlas parcellation,^37^ resulting in 43 ROIs for AT and 49 for PM (Supplementary Tables 1-2). Identical statistical analyses were then performed on the average connectivity of each ROI. Results were considered significant after Holm-Bonferroni’s controlling procedure to account for multiple tests (*pcorr* < 0.05). Within AT, mixed models revealed that functional connectivity increased with age in anterior temporal regions situated in the anterior hippocampus, lateral entorhinal cortex, planum temporal, temporal pole, fusiform, and inferior and middle temporal gyri. In contrast, extra temporal regions located in the superior parietal lobule, postcentral and inferior frontal gyri or posterior regions of the fusiform and temporal gyrus showed decreased connectivity. Within PM, only reduced functional connectivity was significantly observed with age (see Fig. 2 and Supplementary Tables 3-4).

**Figure 2.**
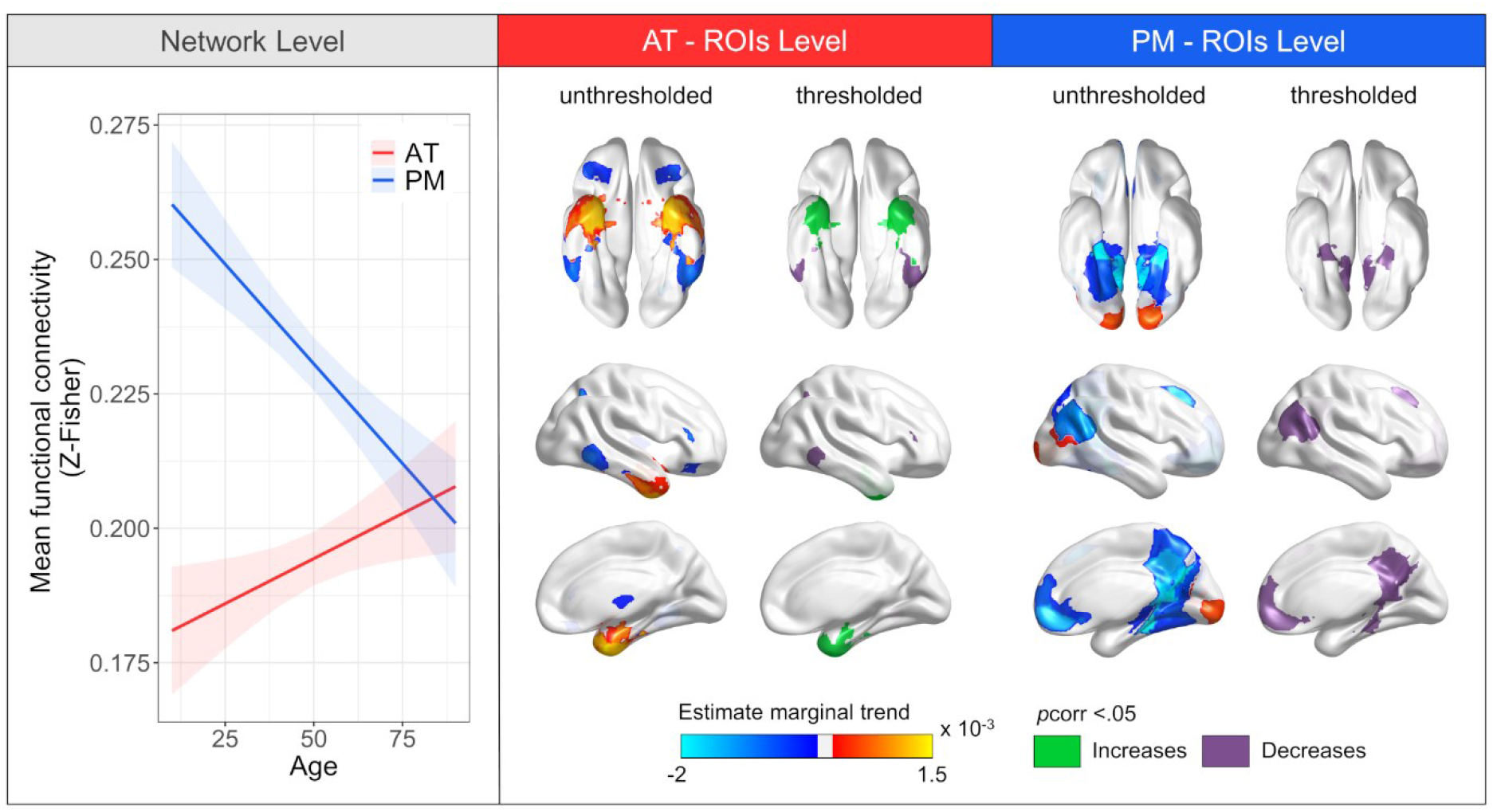
Age-related functional changes in the anterior-temporal (AT) and posterior-medial (PM) networks across the adult lifespan. The network level panel represents age-specific effects on AT and PM global functional connectivity. Connectivity of both networks was significantly related to age (*p* < 0.05). Regression lines indicate model-derived estimates. Shaded areas represent a 95% confidence interval. The ROIs level panel represents age-specific effects on mean functional connectivity within AT and PM regions of interest (ROIs). Unthresholded maps reflect model-derived estimates for all corresponding analyses, including nonsignificant ones. Thresholded maps represent ROIs for which the *p*-value associated to the age term survived to Holm-Bonferroni adjustment for multiple testing (*p_corr_* < 0.05). Green indicates positive associations between age and functional connectivity and purple indicates negative associations.

### Associations with AD neuroimaging biomarkers and clinical manifestations

Baseline data from participants over the age of 60, with and without cognitive impairment, were used to assess AT and PM functional changes across AD stages. Based on their clinical diagnosis and Aβ status, groups were compared using ANCOVA adjusted for age, sex and education. Results revealed differences between groups for AT (*F* = 3.3, *p* < 0.05) but not for PM (*F* = 0.9, *p* = 0.5), such that Aβ-positive patients with MCI or AD-dementia showed greater AT functional connectivity compared to Aβ- negative cognitively unimpaired older adults (*t* = -2.5, *p* < 0.05 and *t* = -2.3, *p* < 0.05, respectively), while Aβ-positive and Aβ-negative cognitively unimpaired older adults or Aβ-positive MCI and AD-demented patients did not differ significantly (see Fig 3, n = 115). Using longitudinal data, linear mixed models did not show significant interactions between time and group for either AT (*F* = 1, *p* = 0.4) or PM (*F* = 1.5, *p* = 0.2), suggesting that functional changes over time did not significantly differ between patient and control groups (Fig. 3, n = 115, data = 240).

**Figure 3.**
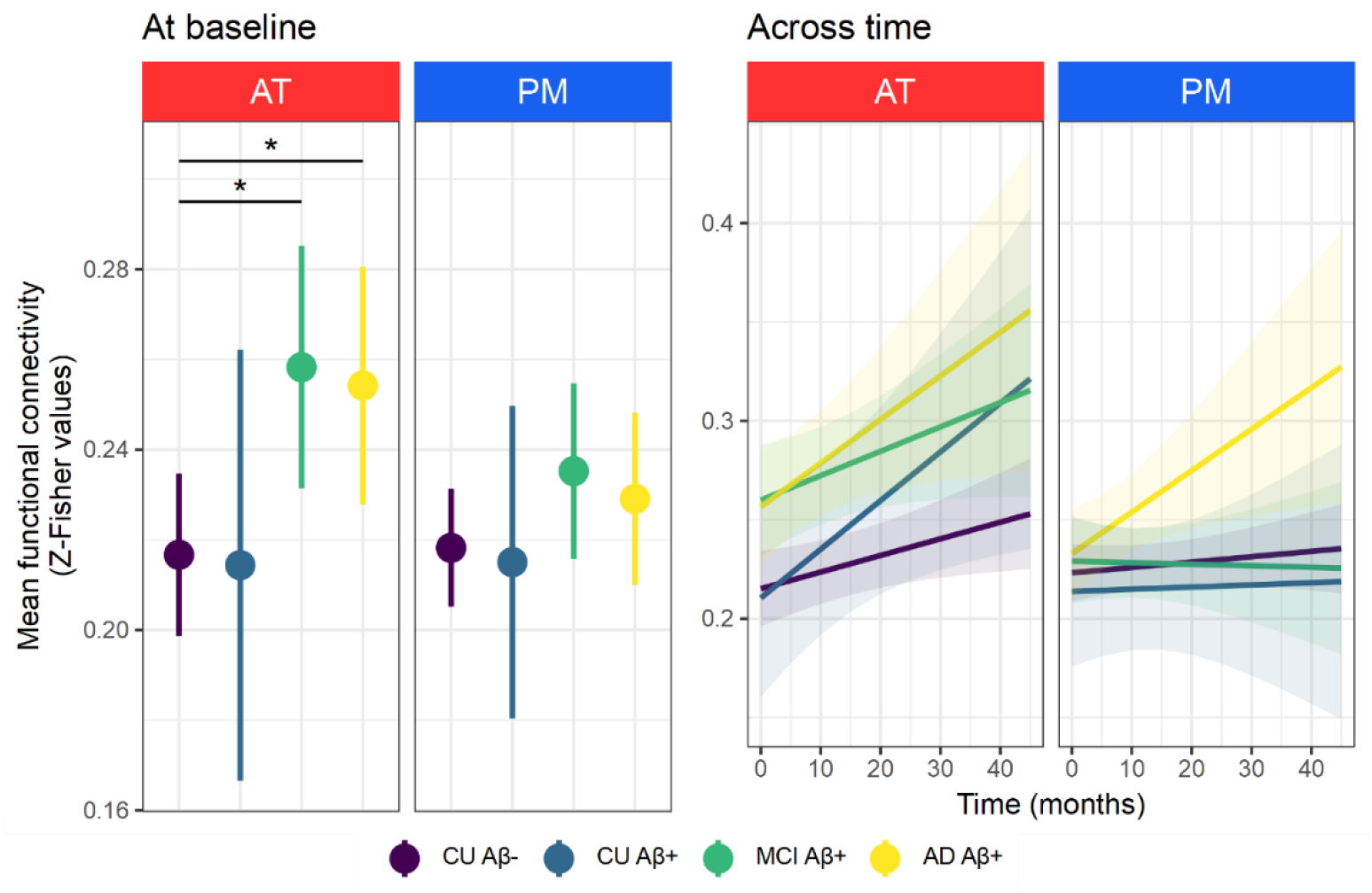
Group differences in functional connectivity within the anterior-temporal (AT) and posterior-medial (PM) networks in older adults, from healthy to AD-demented. Only AT connectivity differed by group at baseline (*p* < 0.05). Point ranges indicate model-derived estimates with 95% confidence intervals. \**p* < 0.05 as results of post-hoc t-tests. No significant time by group interaction was found for AT or PM using longitudinal data. Regression lines indicate model-derived estimates trend. Shaded areas represent 95% confidence intervals.

Associations with AD signature neuroimaging and cognitive markers were further explored using separate linear mixed models adjusted for age, sex, education and time, applied to the same population (10 tests). Neuroimaging biomarkers were derived from PET and MRI scans using the following methods: standardized uptake value ratios (SUVR) for amyloid uptake and glucose metabolism were extracted from PET images in previously defined AD-sensitive regions,^38^ and hippocampal volume was calculated from T1-weighted MRI using the Automatic Segmentation of Hippocampal Subfields (ASHS) software^34^ (see <u>Methods</u>). Global cognition was assessed using the Mini-Mental State Examination (MMSE) and Mattis Dementia Rating Scale (MDRS). Results showed that AT but not PM connectivity was associated with all available AD features (Fig. 4A, all *p*corr < 0.01 and all *p*corr > 0.1, for AT and PM respectively). Specifically, higher functional connectivity in AT was associated with higher amyloid uptake (*F* = 17.3, *p*corr < 0.001, 95% CI = 8.4e^-2^, 2.4e^-1^, n = 114, data = 206), lower glucose metabolism in AD-sensitive regions (*F* = 28.2, *p*corr < 0.001, 95% CI = -1.4e^-1^, -6.4e^-2^, n = 112, data = 218), smaller hippocampal volume (*F* = 12.0, *p*corr < 0.01, 95% CI = -1.1e^-1^, -3.0e^-2^, n = 114, data = 237) and worse global cognition (*F* = 13.4, *p*corr < 0.01, 95% CI = -7.2e^-3^, -2.2e^-3^, n = 114, data = 237 for the MMSE; and *F* = 15.2, *p*corr < 0.01, 95% CI = -2.8e^−3^, -9.0e^−4^, n = 113, data = 224 for the MDRS). Exploratory ROI-based investigations were also conducted to capture regional differences within the networks (Supplementary Tables 5-6). Results evidenced that anterior temporal regions of AT, located in the anterior hippocampus, lateral entorhinal cortex, temporal pole, planum temporal, fusiform, and inferior, middle and superior temporal gyri, were preferentially associated with all features of interest: amyloid uptake (number of significant ROIs [n] = 11, less to most significant ROI: *F* = 8.8 to 24.6, *p*corr < 0.05 to 0.001), glucose metabolism (n = 18, *F* = 8.5 to 31.4, *p*corr < 0.05 to 0.001), hippocampal volume (n = 8, *F* = 9.6 to 21.9, *p*corr < 0.05 to 0.001), MMSE (n = 7, *F* = 11 to 28.1, *p*corr < 0.05 to 0.01), MDRS (n = 9, *F* = 9.2 to 27.8, *p*corr < 0.05 to 0.001) (see Fig. 4B and Supplementary Tables 7-11). Interestingly, higher amyloid uptake (n = 6, *F* = 11 to 20, *p*corr < 0.05 to 0.001), lower glucose metabolism (n = 5, *F* = 11.4 to 17.3, *p*corr < 0.05 to 0.001) and lower MMSE score (n = 6, *F* = 11.3 to 18.9, *p*corr < 0.05 to 0.001) were also associated with higher functional connectivity in predominantly temporal ROIs of PM, located in the medial entorhinal cortex, pulvinar thalami, and medial occipito-temporal, inferior temporal and fusiform gyri (Fig. 4B; Supplementary Tables 12-16).

**Figure 4.**
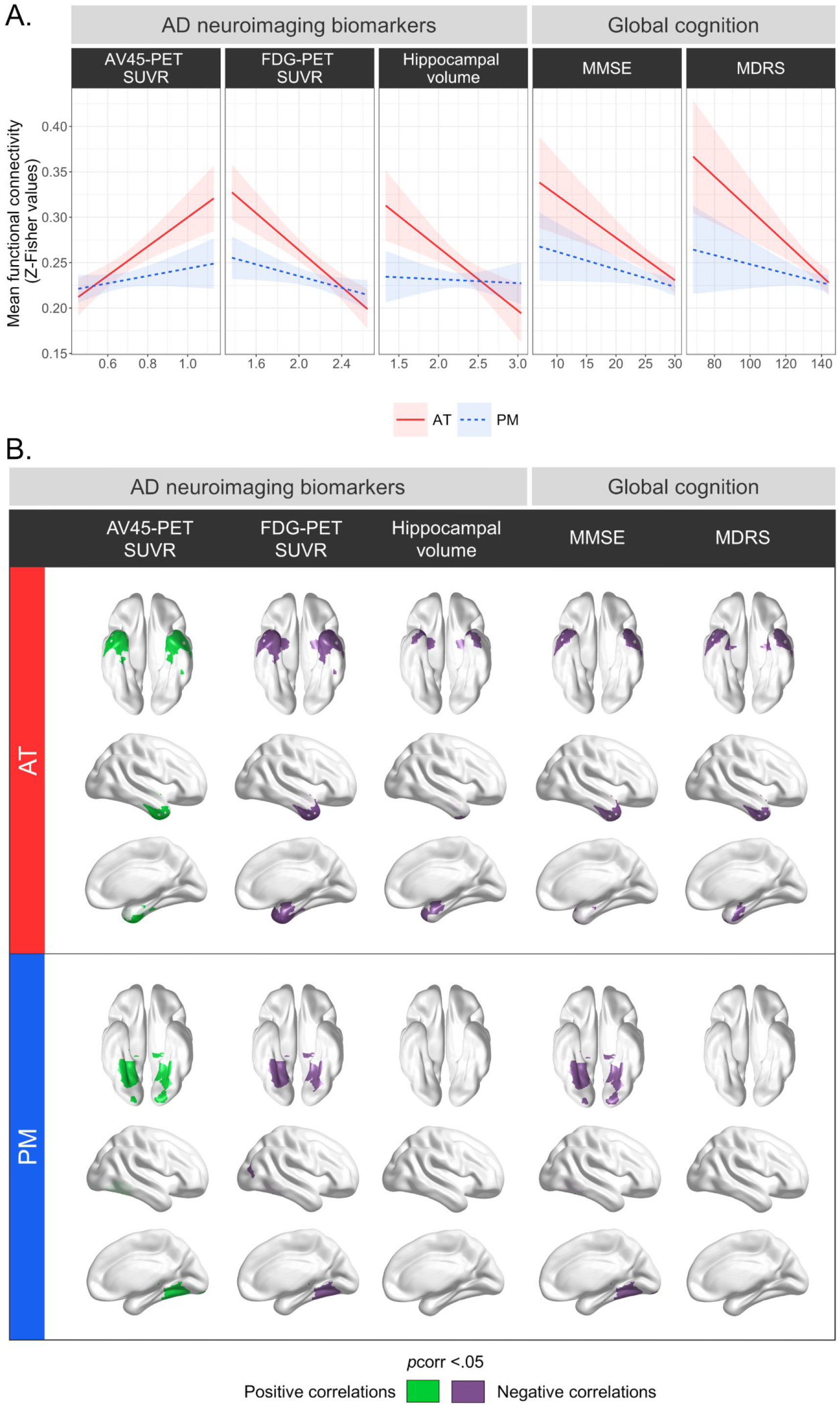
Associations between functional connectivity within the anterior-temporal (AT) and posterior-medial (PM) networks and neuroimaging biomarkers and clinical manifestations of Alzheimer’s disease (AD). A. Relationships between AT and PM global functional connectivity and AD biomarkers and clinical scores. Filled lines represent significant associations and dashed lines nonsignificant ones (*p_corr_* > 0.05). **B.** Brain maps highlighting Brainnetome-based regions of interest (ROIs) for which the *p*-value associated to the model term of interest survived to Holm-Bonferroni correction (*p_corr_* > 0.05). Green indicates positive correlations between the feature of interest and functional connectivity and purple indicates negative correlations. MDRS: Mattis Dementia Rating Scale, MMSE: Mini-Mental State Examination.

### Dementia onset in MCI converters

Within up to 7 years of clinical follow-up, 18 of the 26 Aβ-positive MCI patients evolved to AD-dementia (69%). Time between fMRI exam and dementia onset was calculated for all scans among these participants, and its association with AT and PM functional integrity was assessed using linear mixed models adjusted for age, sex and education. Results indicated that higher AT functional connectivity was associated with faster progression from MCI to AD-dementia (*F* = 6.7, *p* < 0.05, 95% CI = -2.5e^−2^, -1.8e^−3^), while no relationship was found for PM (*F* = 1.6, *p* = 0.2) (see Fig. 5, n = 18, data = 76).

**Figure 5.**
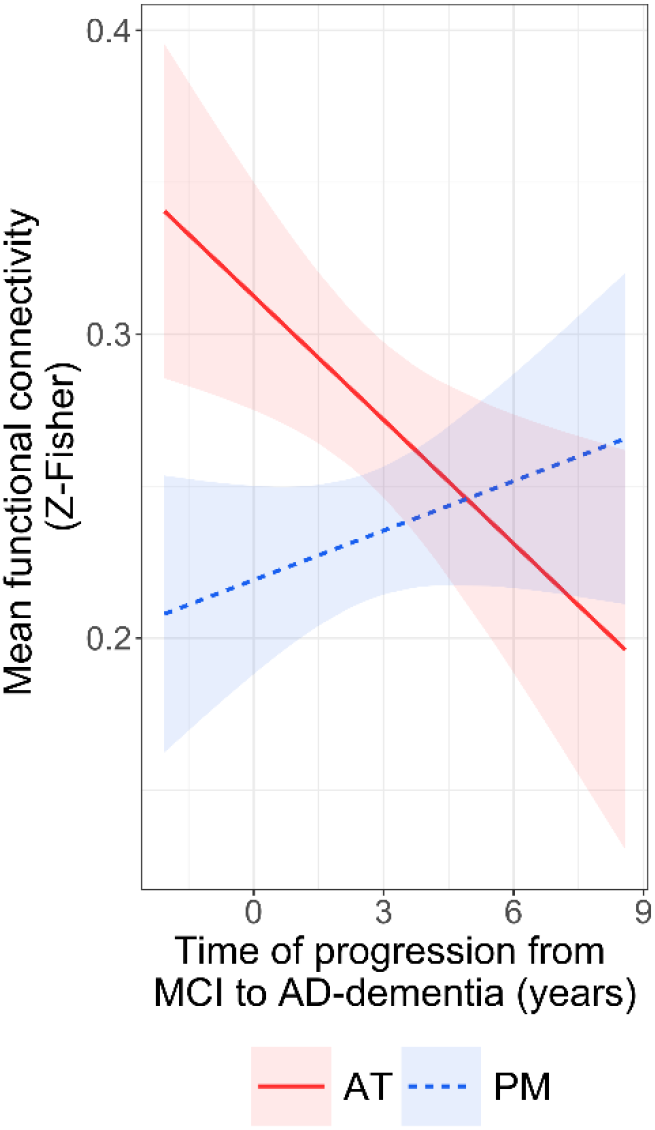
Associations between functional connectivity in the anterior-temporal (AT) and posterior-medial (PM) networks and delay to AD-dementia onset in patients with mild cognitive impairment (MCI). Regression lines indicate model-derived estimates. Shaded areas represent a 95% confidence interval. Filled line represents a significant association and dashed line a nonsignificant one (*p* > 0.05).

### Functional trajectories from physiological to AD-related pathological aging

Changes associated with age and AD were examined together along a common continuum encompassing the whole sample, ranging from young cognitively unimpaired to older demented adults. Generalized additive models applied to baseline data showed that AT functional connectivity increased consistently across the whole spectrum, covering the entire adult lifespan and the entire cognitive continuum from normal to dementia (*F* = 36.0, *p* < 0.001, edf = 1). In contrast, PM connectivity decreased during normal aging and slightly increased across the AD continuum (*F* = 10.1, *p* < 0.001, edf = 2.3) (see Fig. 6, n=259). Trajectories were similar using both sets of masks-derived from cognitively unimpaired young or older adults (see Supplementary Table 17). Longitudinal changes across the whole sample spectrum were evaluated using linear mixed models revealing a significant interaction between group and time for AT (*F* = 4.6, *p* < 0.05) but not for PM (*F* = 1.0, *p* = 0.3), suggesting that AT functional connectivity increased more rapidly over time when progressing from physiological to pathological aging (Fig. 6, n = 260, data = 498).

**Figure 6.**
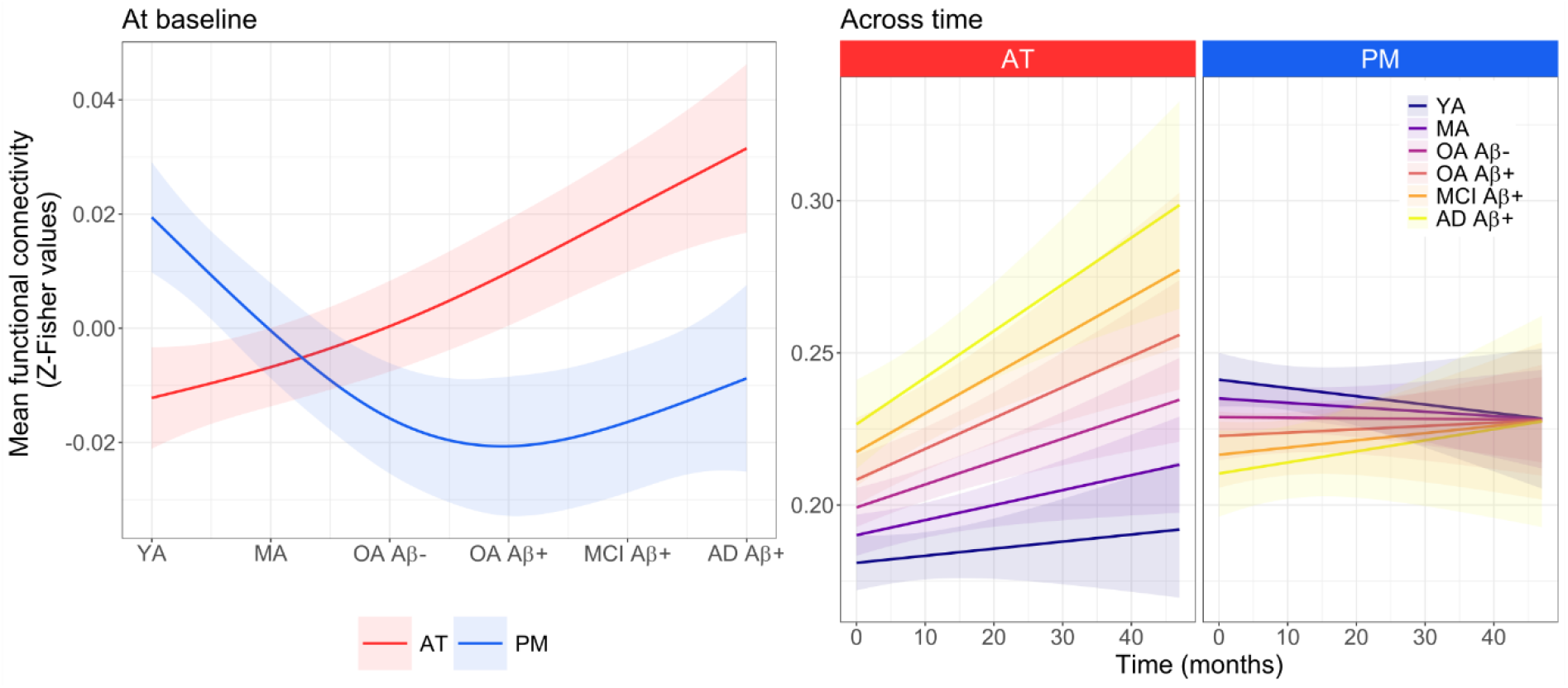
Functional changes in the anterior-temporal (AT) and posterior-medial (PM) networks from physiological to pathological aging due to Alzheimer’s disease (AD). Baseline connectivity of both networks was significantly related to group (*p* < 0.001). Functional connectivity in AT, but not in PM, increased significantly across the whole sample spectrum over time (*p* < 0.05). Continuous lines indicate model-derived estimates. Shaded areas represent a 95% confidence interval. AD: Alzheimer’s disease, MA: middle-aged adults (40-60), MCI: mild cognitive impairment, OA: older adults (>60), YA: young adults (19-39).

Lastly, all analyses were repeated including both AT and PM in the same model to test if they differ significantly in their changes during physiological and pathological aging. All models showed a significant interaction, suggesting substantial differences between the networks. Post hoc analyses evidenced similar findings than those obtained with separate models for AT and PM. In AT, connectivity was positively associated with age over the adult lifespan, and with cerebral amyloid burden and diagnosis severity across the AD continuum, and negatively associated with cerebral glucose metabolism in AD regions, hippocampal volume, global cognitive performance and delay to dementia onset across the AD continuum. In PM, connectivity was negatively associated with age over the adult lifespan and showed no significant association with AD features (<u>see</u> Supplementary Table 18).

## Discussion

In this paper, we present fundamental insights into the functional dynamics of medial temporal lobe networks using longitudinal data spanning physiological aging to the full range of AD impairments. Our study emphasizes the dissociable effects of age and AD, and argue for the relevance of medial temporal lobe connectivity in detecting and monitoring the progression of AD through a collection of converging evidence pertaining to both biological and clinical aspects.

Our findings indicate that AD differentially affects medial temporal lobe networks, with specific disruption of AT over PM. Importantly, higher amyloid burden and lower glucose metabolism in AD signature regions, smaller hippocampal volume, poorer cognitive performance, and more advanced clinical diagnosis were consistently linked to increased AT connectivity. While prior research on AT dysfunction in AD led to inconsistent findings, our collection of new evidences advocates that AT hyperconnectivity is instrumental for AD pathophysiology. This substantiates previous findings describing greater functional connectivity within AT in MCI or AD patients compared to controls.^25–27,39^ Possibly, the process of AT hyperconnectivity is initiated by early tau accumulation in the hub area,^18^ where neurons may be more vulnerable to adverse conditions that facilitate aggregates to emerge.^40^ This hyperconnectivity, in turn, may stimulate the secretion of tau locally.^41^ In the presence of Aβ pathology, disruption in network activity may result in connectivity-mediated interaction between cortical Aβ and localized tau. This remote interaction induces tau to undergo biophysical changes, which drives it to spread into connected temporal regions. Subsequent local interaction with pre-existing Aβ may catalyse widespread tau propagation into Aβ-positive connected regions,^20^ resulting in increased neuron toxicity. Ultimately, this may lead to hypometabolism and neurodegeneration,^42^ giving way to cognitive impairment. This metabolic imbalance may also boost irregular hyperconnectivity, creating a vicious cycle of AD progression through a positive feedback loop.^43^ Such mutually dependent mechanisms could account for prior evidence suggesting that abnormal cell firing in the medial temporal lobe indicates impending mortality.^44^ Yet, further insight will be required to accurately determine the causal relationships underlying this circuit dysfunction. Since abnormal calcium signalling in astrocytes is known to affect neurovascular coupling and could lead to neuronal hyperactivity,^45,46^ astrocytic deficiency or inflammation might be further explored as possible explanations for aberrant AT activity in AD. Prognostically, we deliver compelling evidence that AT hyperconnectivity indicates hastened disease advancement, as increased AT functional connectivity was associated with faster dementia onset in MCI patients who progressed to AD-dementia. Such findings represent a pioneering investigation into the clinical course of patients regarding medial temporal lobe networks’ dynamics, and align with the prior identification of hippocampal hyperactivity in individuals at risk for AD.^47^ At the network level, no correlation were found between PM functional connectivity and any features associated with AD pathology or clinical staging, although previous studies have robustly documented reduced connectivity within PM in AD patients. We explain these discrepancies by our methodological choice to limit our analyses to the connectivity of the parahippocampal cortex. In fact, reports of reduced PM functional connectivity in AD often refer to disconnections with anterior regions outside our mask or disconnections between PM components outside the medial temporal lobe,^25,26,48^ which are not captured by our connectivity maps. Further work not focusing on the hub region may shed light on such disconnections and their links with pathological and clinical outcomes. This drawback, however, yield more easily deployable neuroimaging biomarkers for clinical use by providing comparable measures across analyses. Interestingly, ROI level analyses within PM indicated that AD features were more likely to be associated with increased connectivity in temporal regions of PM. These regions encompassed the entorhinal cortex, fusiform and inferior temporal gyrus. Overall, our findings support earlier research indicating medial temporal lobe hyperexcitability in AD,^16,48,49^ and strengthen animal evidences that hippocampal hyperactivity, rather than hypoactivity, is the primary neuronal dysfunction.^50^

To our knowledge, our study is the first to demonstrate that AT and PM undergo distinct aging processes in a large sample spanning the entire adulthood, with longitudinal follow-up. We emphasized that PM functional connectivity decreased throughout adult life, which is consistent with the recent evidence of an age-related decline in parahippocampal connectivity.^33^ Conversely, the anterior temporal regions of AT, which are particularly affected by early accumulation of neurofibrillary tangles,^5^ exhibited increased functional connectivity with age. This hyperconnectivity within AT may therefore indicate primary age-related tauopathy, in light of connectome-based models.^16,51,52^ In contrast, the frontal, parietal and posterior temporal regions of AT displayed reduced connectivity. Such heterogeneity within AT may account for previous inconsistencies in findings reporting either increased or reduced medial temporal lobe connectivity with older age.^30–32,48^ Interestingly, age effects were more pronounced for PM than for AT, which substantiate the greater functional vulnerability of posterior brain regions to aging.^31,33^ This particular sensitivity may explain cognitive impairment experienced in old age, which preferentially affect associative memory and spatial cognition, which are predominantly supported by PM components.^6,7,53,54^ Altogether, our results support the previous observation of an age-related decoupling between regions of the default mode networks and the medial temporal lobe, resulting in functional isolation of the hippocampus and its surrounding regions.^30,55–57^

Our data-driven analyses, using generalized additive models across the entire sample, clearly illustrated the specific effects of physiological and pathological aging on functional connectivity in the medial temporal lobe networks. While both advanced age and AD led to increased connectivity within AT, this process was accelerated across the AD continuum. In addition to baseline differences, AT connectivity also increased more rapidly over time, suggesting that AT hyperconnectivity-related consequences worsen with AD progression. Prior animal research suggests that early stages of AD are characterized by neuronal hyperactivity, followed by functional silencing of neurons later on.^58^ A recent neuroimaging study focusing on medial temporal lobe functional connectivity substantiated this claim by demonstrating greater connectivity within AT during preclinical phases but lower connectivity in advanced stages of AD.^33^ Despite non-a priori inferences, our analyses proffer no evidence for such an inverted U-shaped trajectory across AD stages. However, only mild to moderate demented patients were included in our study, so further consideration of severe dementia may reveal late AT outcomes. In contrast, PM underwent different changes from physiological to pathological aging. Consistent with our previous findings, reduced connectivity within PM was found to specifically reflect age-related processes. In addition, slight increased PM connectivity was observed across the AD continuum, potentially accounting for the AD-featured hyperconnectivity within temporal ROIs of PM. These findings align with the theoretical framework that early pathological changes are marked by weakening of distal connections and strengthening of proximal connections,^16^ thereby providing preliminary evidence for network dedifferentiation throughout AD progression. Based on prior work suggesting reduced network specialization due to AD pathology within AT and PM in older adults,^59^ we hypothesize that the medial temporal lobe networks function independently in early adulthood and undergo dedifferentiation processes when AD manifests.

The main strengths of this study include the monocentric dataset with available longitudinal (f)MRI, amyloid- and FDG-PET, clinical and cognitive data, which allowed us to estimate individual risk for AD progression. In addition, we opted for a fine methodology reaching for individual-level optimal seed to ensure networks accuracy and replicability. Another key point is the development of easier-to-interpret fMRI outcomes, thought out as a standardized index to provide a personalized prognosis that reliably estimates the risk of future cognitive decline. Nevertheless, our design only counts a limited follow-up and small sample size. Specifically, a larger pool of Aβ- positive cognitively unimpaired older adults may be crucial to draw conclusions about early diagnosis, as only 8 participants were classified as preclinical AD. This is particularly important because other studies have already pointed to differences at this stage^26,48^ and our analyses of AD neuroimaging features endorse the importance of AT functional connectivity in early AD-related brain changes. Longer follow-up may also provide more sensitivity to our analyses, especially with respect to changes over time that may be undetectable with a limited number of scans.

In conclusion, our longitudinal study yields important findings into the functional dynamics of medial temporal lobe networks during physiological to pathological aging and its links with AD-associated clinical and pathological hallmarks. Over and above prior research, our results converge to emphasize that medial temporal lobe hyperconnectivity is not just a disease feature, but a fundamental aspect of AD process and likely serves as a catalyst for its progression. Uncovering such mechanisms helped us to shed light on AD pathophysiology and holds promise for the development of early diagnostic tools and therapeutic interventions targeting these specific network alterations.

## Methods

Unless otherwise noted, all data analyses were conducted and all figures were created using BrainNet Viewer version 1.6 and R version 4.2.0, mostly using the lmerTest, gamm4, emmeans and ggplot2 libraries.

### Participants

Two hundred sixty-seven native French-speaking participants enrolled in the *Imagerie Multimodale de la Maladie d’Alzheimer à un stade Précoce* (IMAP) study (Caen, France) between 2008 and 2016 were included in the present study. All were right-handed with 7 years of education minimum, and no history of alcoholism, drug abuse, head trauma, neurological or psychiatric disorder. The IMAP study was approved by a regional ethics committee (Comité de Protection des Personnes Nord-Ouest III) and was registered with ClinicalTrials.gov (number: NCT01638949). All participants gave written informed consent to the study before the investigation.

Two hundred and fifteen cognitively unimpaired adults aged from 19 to 85 were recruited from the community using flyers and advertisements in local newspapers. Their cognitive performances were within the normal range for their age and education level, and they did not express memory complaints. Fifty-two patients were recruited from local memory clinics and selected according to internationally agreed criteria. Amnesic patients with MCI satisfied Petersen’s criteria^60^ (n = 26) and patients with AD-dementia fulfilled the standard National Institute of Neurological and Communicative Disorders and Stroke and the Alzheimer’s Disease and Related Disorders Association (NINCDS-ADRDA) clinical criteria for mild to moderate probable AD^61^ (n = 26). Clinical diagnosis was assigned by consensus under the supervision of a senior neurologist (V.d.l.S.).

Participants were followed up to 4 years with 1-3 assessments of neuroimaging and cognitive data (± 18 months apart). All participants older than 60 years underwent an F^18^-AV45 PET exam to assess amyloid-β deposition. In cognitively unimpaired older adults, the proportion of Aβ-positive was 12.5% (8/64). The positivity threshold was calculated as the 99.9th percentile of the SUVR distribution from 45 participants younger than 40, corresponding to a SUVR of 1.31. Aβ status was determined from baseline scan for most of our population, but when no PET scan was available at baseline and the SUVR from subsequent scans was below the threshold, individuals were considered Aβ-negative (n = 21).

Patients included as MCI were clinically followed until December 2022. From this sample, 69% (18/26) evolved from MCI to AD-dementia within 7 years. Dementia diagnosis was posited in diagnostic commission by a senior neurologist (V.d.l.S) according to standard clinical criteria and based on feedback from the patient clinicians.

### Neuroimaging data acquisition and pre-processing

Participants were repeatedly scanned with the same MRI and PET scanners at the Cyceron Center (Caen, France): a Philips Achieva 3.0 T scanner and a Discovery RX VCT 64 PET-CT device (General Electric Healthcare), respectively.

T1-weighted anatomical images were acquired using a 3D fast-field echo sequence (3D- T1-FFE sagittal; SENSE factor = 2; repetition time = 20 ms; echo time = 4.6 ms; flip angle = 10°; 180 slices with no gap; slice thickness = 1 mm; field of view = 256 × 256 mm^2^; in-plane resolution = 1 × 1 mm^2^). Non-Echo Planar Imaging (EPI) T2* volumes (2D-T2*-FFE axial; SENSE factor = 2; repetition time = 3514 ms; echo time = 30 ms; flip angle = 90°; 70 slices with no gap; slice thickness = 2 mm; field of view = 256 x 256 mm²; in-plane resolution = 2 x 2 mm²) were additionally obtained for fMRI pre- processing steps. Resting-state functional MRI scans were obtained using an interleaved 2D T2* SENSE EPI sequence designed to reduce geometric distortions (2D-T2*-FFE- EPI axial, SENSE = 2; repetition time = 2382 ms; echo time = 30 ms; flip angle = 80°; 42 slices with no gap; slice thickness = 2.8 mm; field of view = 224 × 224 mm^2^; in plane resolution = 2.8 × 2.8 mm^2^; 280 volumes, acquisition time = 11.5 min). Of note, 11 scans from cognitively unimpaired adults younger than 60 years were acquired using a slightly different sequence (2D-T2*-FFE-EPI axial, SENSE = 2; repetition time = 2200 ms; echo time = 35 ms; flip angle = 80°; 35 slices with no gap; slice thickness = 3.5 mm; field of view = 224 × 224 mm^2^; in plane resolution = 3.5 × 3.5 mm^2^; 300 volumes, acquisition time = 11.11 min). Difference between sequences was controlled in statistical analyses. During acquisition, participants wore earplugs and their heads were stabilized with foam pads to minimize head motion. The light in the scanner room was turned off and participants were asked to relax by lying down and closing their eyes, without falling asleep. A short debriefing after the acquisition ensured that participants had no difficulty staying awake and that nothing distracting them. EPI data were first checked visually for movement, lesions, abnormalities, head misplacement and signal loss (especially as temporal regions may be particularly affected). The TSDiffana routine (http://imaging.mrc-cbu.cam.ac.uk/imaging/DataDiagnostics) was applied to functional raw volume to identify artefacts. Data showing evidence for significant movements (>3 mm translation or 1.5° rotation) associated to image artefacts and/or an abnormal variance distribution were considered for exclusion. Pre-processing steps included slice timing correction, realignment to the first volume, rigid coregistration onto T1 images and spatial normalization using SPM12. Individual T1-to-EPI coregistration was performed as followed: 1) mean EPI onto non-EPI T2*, 2) non-EPI T2* onto T2, 3) T2 onto T1. Spatial normalization within the native space was devised to reduce geometrical distortion^62^. Mean EPI image was warped to roughly match non-EPI T2* volume. Then, EPI-to-non-EPI warping parameters were combined to normalization parameters obtained from previously segmented T1 images onto the Montreal Neurological Institute (MNI) template and applied to the coregistered T1, EPI, and non-EPI T2* volumes. Resulting images were smoothed with a 4 mm full width at half maximum Gaussian kernel and then band pass filtered (0.01-0.08Hz) using AFNI.

Both ^18^F-fluorodeoxyglucose (FDG) and florbetapir (AV45) PET scans were acquired (resolution = 3.76 x 3.76 x 4.9 mm; field of view = 157 mm; voxel size = 1.95 x 1.95 x 3.27 mm). A transmission scan was performed for attenuation correction before the PET acquisition. For FDG-PET, participants fasted for at least 6 hours before scanning and remained in a quiet, dark environment for 30 minutes before injection of the radiotracer. Fifty minutes after the intravenous injection of 5.4 mCi of ^18^F-FDG, a 10 minutes PET acquisition began. For AV45-PET, data were locally reconstructed into 4 x 5 min frames for the 50 to 70-minute interval after injection of 4 MBq/kg of florbetapir. PET data were coregistered onto their corresponding T1-weighted MRI and normalized to the MNI space using the parameters from the T1 segmentation procedure. Resulting images were quantitatively normalized using the cerebellum or white matter uptake as reference depending on analyses (i.e. cross-sectional vs longitudinal, respectively). As Aβ-status were determined from participant first scan corresponding to their clinical diagnosis at baseline, global SUVRs of neocortical amyloid were extracted from cerebellum-scaled AV45-PET images. In contrast, white matter-scaled AV45- and FDG-PET images were used to extract SUVRs reflecting amyloid uptake and glucose metabolism. Extractions were done in AD-sensitive regions (i.e. brain regions exhibiting high amyloid burden or severe hypometabolism in AD)^38^ within a subject-specific grey matter mask derived from T1-weighted MRI.

### Medial temporal lobe regions segmentation

The hippocampus, perirhinal and parahippocampal cortices were automatically segmented on T1-weighted scans using the Automatic Segmentation of Hippocampal Subfields (ASHS) software^63^ (atlas: ashsT1_atlas_upennpmc_07202018). The standard ASHS-T1 pipeline was applied in participants with a single neuroimaging exam, while the Longitudinal Automatic Segmentation of Hippocampal Subfields (LASHiS) pipeline^64^ was used for participants with multiples scans. As previously described, each segmentation was visually inspected.^4^ Failed segmentations were manually edited when doable, or excluded. Left and right structures were merged to limit the number of analyses. Hippocampal volume was calculated from the output segmentation combining the anterior and posterior hippocampus and then normalized by the total intracranial volume (TIV) to account for inter-individual variability in head size (normalized volume = raw volume / [TIV x 1000]). The perirhinal -consisting of Brodmann areas 35 and 36- and parahippocampal cortices were normalized to MNI space and used as seeds for the connectivity analyses.

### Functional connectivity assessment

Resting-state fMRI scans were processed using SPM12 and the MarsBaR toolbox (version 0.44), following the steps depicted in Fig. 1. Positive correlations were assessed between the mean time course in ASHS-T1-derived seeds (i.e. the perirhinal and parahippocampal cortices) and the time course of each grey matter voxel, resulting in 2 grey matter connectivity maps for each fMRI scan. The low-frequency drifts were removed for each seed. Mean time courses in white matter, cerebrospinal fluid, whole brain, their derivatives, and 6 movement parameters generated from realignment of head motion were regressed out to remove potential sources of spurious variance. A Fisher z transformation and a 6.3 mm FWHM smoothing were finally applied to the individual connectivity maps. Two separate one-sample permutation t-tests were performed on smoothed perirhinal and parahippocampal maps using the FSL randomise command with TFCE and FWE correction to retain only regions significantly connected to the seed (*p*corr < 0.05). Simultaneously, perirhinal to parahippocampal maps (and vice versa) were compared using paired permutation t-tests with identical settings to highlight regions specific to AT (regions more strongly connected to the perirhinal than to the parahippocampal seed) and to PM (regions more strongly connected to the parahippocampal than to the perirhinal seed). Output t-maps were thresholded at *p*corr < 0.05 to provide 95% confidence of no false positives, and then binarized to define the AT and PM masks. Masks consisted of the overlap between the output binarized maps from the one-sample and two-sample paired t-tests (perirhinal one-sample + perirhinal > parahippocampal paired t-tests for AT, and parahippocampal one-sample + parahippocampal > perirhinal paired t-tests for PM). To obtain intact networks relevant for both age- and AD-related analyses, our t-tests routine was restricted to baseline data from cognitively unimpaired adults under the age of 40 for age-related analyses and from Aβ-negative cognitively unimpaired adults over the age of 60 for AD-related analyses (Fig 1<u>.B</u>). Individual grey matter masks were created for all participants from the corresponding T1 grey segmentation thresholded at 0.5. Group-level AT and PM masks were multiplied by these individual grey matter masks to derive subject-specific AT and PM masks accounting for inter-individual anatomical variability. Seeds were removed from masks to avoid auto-correlations. Mean connectivity within subject-specific AT and PM masks was extracted from unsmoothed perirhinal and parahippocampal maps, respectively, for all available data. The average signal was computed on positive voxels only because negative values were close to 0, probably reflecting noise not biologically meaningful.^65^ To examine regional differences within AT and PM connectivity, masks were split according to the Brainnetome atlas parcellation,^37^ resulting in 43 ROIs for AT and 49 for PM for the masks based on young adults (Supplementary Tables 1-2), and in 24 ROIs for AT and 41 for PM for the masks based on older adults (Supplementary Tables 5-6). Only ROIs including at least 100 voxels were investigated. The average connectivity within each ROI was then calculated as described above.

### Statistical analyses

Unless otherwise stated, all statistical analyses based on longitudinal data were performed using linear mixed models because they handle within-subject observations, missing data and unbalanced designs, such as irregular follow-ups in the current study. Generalized additive (mixed) models were also used, as they allow nonlinear relationships without a priori inferences. Models were fitted to the data with restricted maximum likelihood. Type II or III F-tests -depending on the absence or presence of an interaction term- with Satterthwaite’s method for degrees of freedom approximation were used to assess the significance of fixed effects. When analyses implied multiple testing, results were considered significant after Holm’s family-wise error rate controlling procedure (*p*corr < 0.05).^66^

Age-specific effects were assessed among cognitively unimpaired adults (aged 19-85 at baseline) using the set of masks defined in young adults. Separate models for AT and PM were run. Average connectivity within each network was entered as dependent variable and age as predictor -calculated as the time difference between birth date and fMRI exam. Sex, years of education and fMRI sequence parameters were added as covariates. Random intercepts per participants were computed. Similar models were also conducted at the ROI-level to outline regional differences within the AT and PM networks.

Functional alterations across the AD continuum were investigated in all participants older than 60, including Aβ-negative and Aβ-positive cognitively unimpaired older adults and Aβ-positive MCI and AD patients, using the set of masks defined in older adults. Baseline differences were assessed using ANCOVAs with group as predictor. Changes over time were estimated using linear mixed models with group, time between baseline and follow-up fMRI exams, and their interaction as fixed effect terms, and random intercepts per participants. For both cross-sectional and longitudinal analyses, AT and PM were investigated in separate models adjusted for age, sex and years of education. Pairwise group comparisons were performed as post-hoc using independent t-tests comparing estimated marginal means.

Links with neuroimaging and cognitive features of AD were investigated in the same individuals using mixed models controlling for age, sex, years of education and time between intra-individual exams. Separate models were conducted for each network and each feature of interest (i.e., amyloid uptake, glucose metabolism, hippocampal volume, MMSE and MDRS scores), leading to 10 tests. Random effects included by-participant intercepts. The same associations were also tested at the ROI-level to highlight regional differences within the AT and PM networks.

Time to dementia onset in MCI converters was tested in association with AT and PM functional connectivity in separate models accounting for age, sex and years of education, and including random intercepts per participants.

Changes across the whole sample spectrum, covering the entire adult lifespan and the entire cognitive continuum from no impairment to dementia, were modelled using generalized additive models with group as a smooth term set to k=6, and sex and years of education as linear terms, using baseline data. Linear mixed models with group, time, and their interaction as fixed effect terms were also computed to assess group differences in change over time. Models included sex and education as covariates, and random intercepts per participant. Analyses were repeated using values extracted from both sets of masks (see <u>Results</u> and Supplementary Table 17).

All analyses were also replicated with linear mixed models incorporating both networks in the same model to shed light on interaction effects. These models included the connectivity value as dependant variable and the interaction between networks and variable of interest as predictor, as well as the relevant covariates to each analysis (see <u>Results</u> and Supplementary Table 18).

## Data availability

Dataset used in the present study is available from G.C. upon reasonable request, for the purpose of replicating procedures and results.

## Code availability

The codes used for data analyses in our study can be requested from L.C. and R.d.F.

## Acknowledgements

Authors are grateful to the patients and healthy volunteers who participated in this study. Thanks to L. Barre, JC. Baron, A. Cognet, J. Dayan, B. Desgranges, S. Egret, F. Eustache, M. Fouquet, J. Gonneaud, D. Guilloteau, R. La Joie, V. Lefranc, A. Manrique, F. Mezenge, K. Mevel, J. Mutlu, G. Poisnel, A. Pélerin, C. Tomadesso, A. Quillard, G. Rauchs, C. Schupp, F. Viader, and N. Villain for their contributions in recruitment, cognitive testing and imaging examinations, and the Cyceron MRI-PET staff members for their help with imaging acquisition.

## Author information

### Contributions

G.C. and V.d.l.S. were the principal investigators of the IMAP research protocol. R.d.F. and L.C. conceived the study. B.L. and L.C. processed neuroimaging data. S.D. and AL. T. helped with the data acquisition and quality control. V.d.l.S., M.D. and O.H. provided their expertise on patient’s clinical condition. L.C. was responsible for statistical coding and analysis. R.d.F., G.C. and L.C. interpreted data. L.C. drafted the manuscript in close collaboration with R.d.F. and G.C. All authors contributed substantially to revising the manuscript.

### Corresponding authors

Correspondence to R.d.F.

