## Supplementary for "Medial temporal lobe hyperconnectivity is key to Alzheimer’s disease: Insight from physiological aging to dementia"

### Supplementary information

**<u>Supplementary Table 1.</u> Summary of Brainnetome-based regions of interest (ROIs) included in the anterior-temporal (AT) mask derived from cognitively unimpaired adults under the age of 40.**

| ROI | Brainnetome labeling | Brain view |
|---|---|---|
| BN_AT_IPL_A39c | Inferior parietal lobule – caudal area 39 | 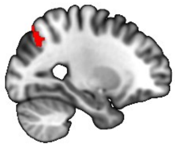 |
| BN_AT_LOcC_lsOccG | Lateral occipital cortex - lateral superior occipital gyrus | 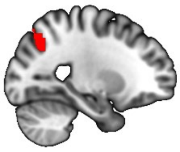 |
| BN_AT_IFG_IFS | Inferior frontal gyrus – inferior frontal sulcus | 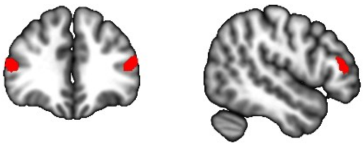 |
| BN_AT_IPL_A39rd | Inferior parietal lobule - rostrodorsal area 39 | 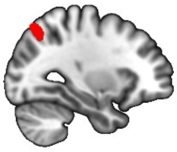 |
| BN_AT_SPL_A7ip | Superior parietal lobule - intraparietal area 7 | 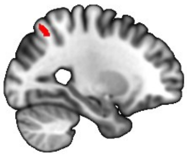 |
| BN_AT_SPL_A7c | Superior parietal lobule - caudal area 7 | 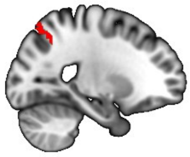 |
| BN_AT_ITG_A37vl | Inferior temporal gyrus - ventrolateral area 37 | 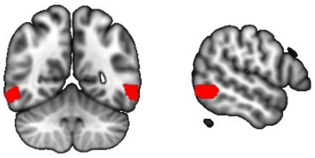 |
| BN_AT_ITG_A37elv | Inferior temporal gyrus - extreme lateroventral area 37 | 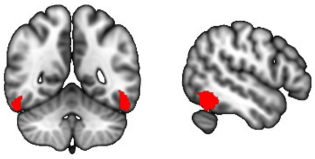 |

| | | |
|---|---|---|
| BN_AT_FuG_A37lv | Fusiform gyrus - lateroventral area 37 | 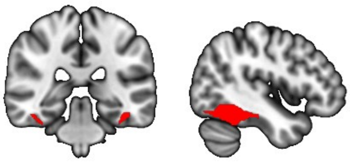 |
| BN_AT_MTG_A37dl | Middle temporal gyrus - dorsolateral area 37 | 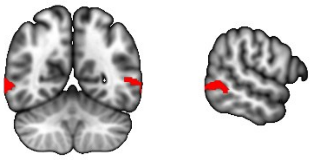 |
| BN_AT_PhG_TL | Parahippocampal gyrus - area TL (lateral posterior parahippocampal gyrus) | 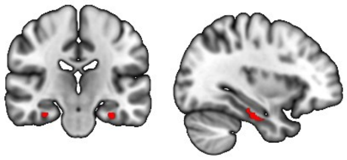 |
| BN_AT_OrG_A12_47o | Orbital gyrus - orbital area 12/47 | 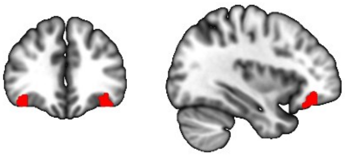 |
| BN_AT_ITG_A20cl | Inferior temporal gyrus - caudolateral of area 20 | 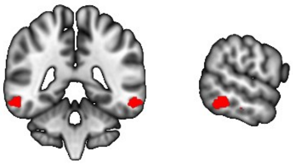 |
| BN_AT_OrG_A11l | Orbital gyrus - lateral area 11 | 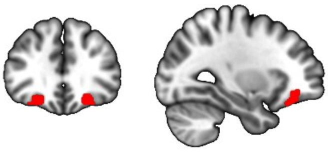 |
| BN_AT_LOcC_V5_MT | Lateral occipital cortex - area V5/MT+ | 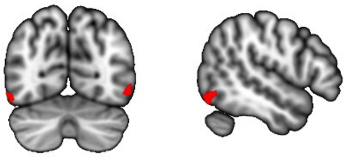 |
| BN_AT_Tha_rTtha | Thalamus - rostral temporal thalamus | 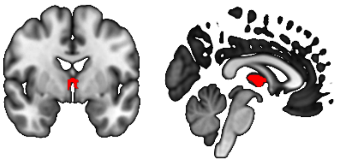 |
| BN_AT_MTG_A21c | Middle temporal gyrus - caudal area 21 | 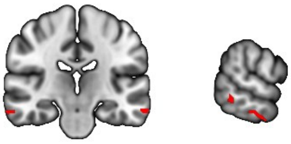 |
| BN_AT_Amyg_mAmyg | Amygdala - medial amygdala | 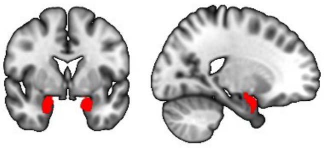 |

| | | |
|---|---|---|
| BN_AT_OrG_A13 | Orbital gyrus – area 13 | 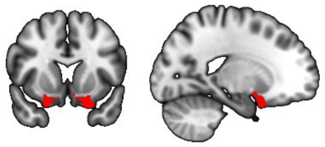 |
| BN_AT_STG_A38l | Superior temporal gyrus - lateral area 38 | 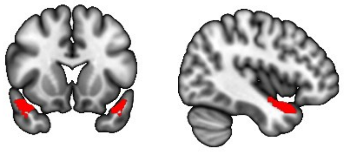 |
| BN_AT_Tha_mPFtha | Thalamus - medial pre-frontal thalamus | 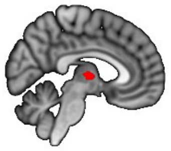 |
| BN_AT_STG_A22r | Superior temporal gyrus - rostral area 22 | 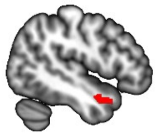 |
| BN_AT_PhG_A35_36c | Parahippocampal gyrus - caudal area 35/36 | 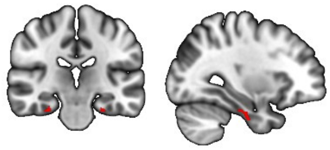 |
| BN_AT_BG_NAC | Basal ganglia – nucleus accumbens | 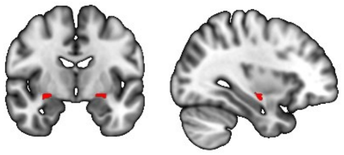 |
| BN_AT_ITG_A20cv | Inferior temporal gyrus - caudoventral of area 20 | 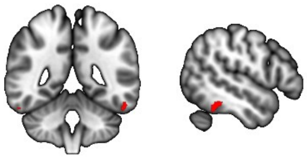 |
| BN_AT_BG_dlPu | Basal ganglia - dorsolateral putamen | 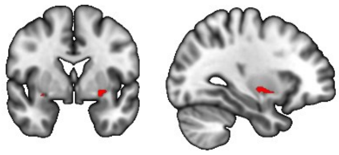 |
| BN_AT_MTG_A21r | Middle temporal gyrus - rostral area 21 | 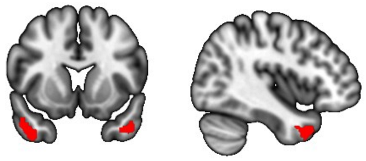 |
| BN_AT_Amyg_lAmyg | Amygdala - lateral amygdala | 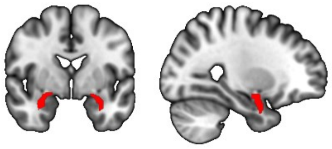 |

| | | |
|---|---|---|
| BN_AT_BG_vmPu | Basal ganglia - ventromedial putamen | 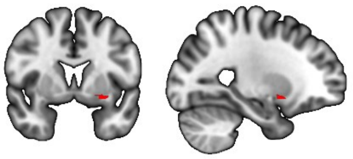 |
| BN_AT_STG_TE10_TE12 | Superior temporal gyrus - TE1.0 and TE1.2 | 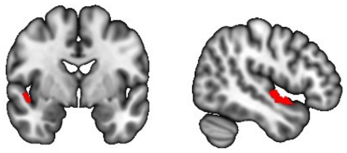 |
| BN_AT_INS_vId_vIg | Insular gyrus - ventral dysgranular and granular insula | 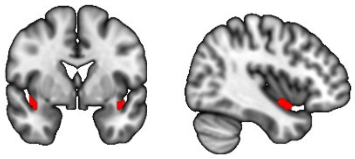 |
| BN_AT_INS_vIa | Insular gyrus - ventral agranular insula | 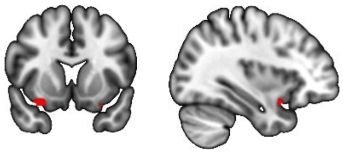 |
| BN_AT_ITG_A20il | Inferior temporal gyrus - intermediate lateral area 20 | 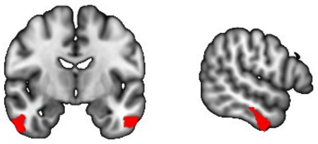 |
| BN_AT_Hipp_rHipp | Hippocampus- rostral hippocampus | 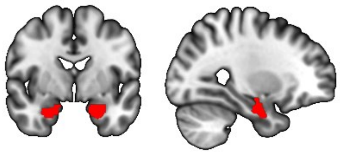 |
| BN_AT_Hipp_cHipp | Hippocampus- caudal hippocampus | 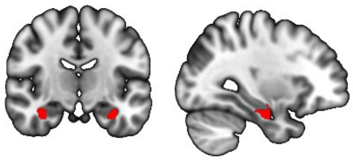 |
| BN_AT_ITG_A20iv | Inferior temporal gyrus - intermediate ventral area 20 | 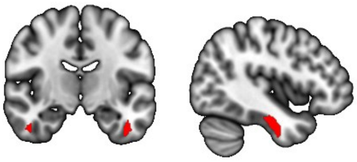 |
| BN_AT_ITG_A20r | Inferior temporal gyrus - rostral area 20 | 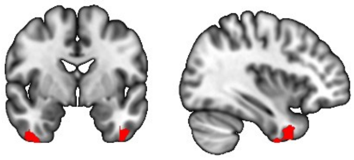 |
| BN_AT_STG_A38m | Superior temporal gyrus - medial area 38 | 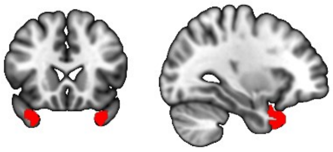 |

| | | |
|---|---|---|
| BN_AT_FuG_A20rv | Fusiform gyrus - rostroventral area 20 | 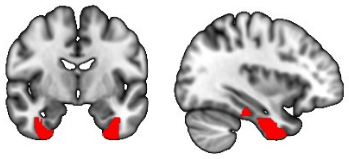 |
| BN_AT_PhG_A35_36r | Parahippocampal gyrus - rostral area 35/36 | 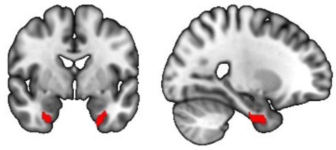 |
| BN_AT_PhG_TI | Parahippocampal gyrus - area TI (temporal agranular insular cortex) | 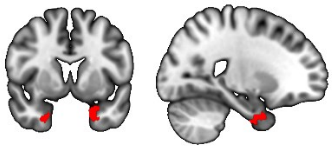 |
| BN_AT_PhG_A28_34 | Parahippocampal gyrus - area 28/34 (entorhinal cortex) | 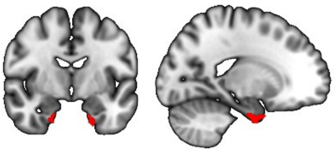 |

**<u>Supplementary Table 2.</u> Summary of Brainnetome-based regions of interest (ROIs) included in the posterior-medial (PM) mask derived from cognitively unimpaired adults under the age of 40.**

| ROI | Brainnetome labeling | Brain view |
|---|---|---|
| BN_PM_PhG_TL | Parahippocampal gyrus - area TL (lateral posterior parahippocampal gyrus) | 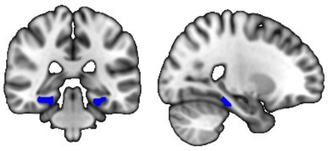 |
| BN_PM_FuG_A20rv | Fusiform gyrus - rostroventral area 20 | 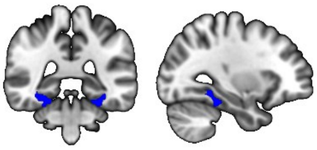 |
| BN_PM_PhG_TH | Parahippocampal gyrus - area TH (medial posterior parahippocampal gyrus) | 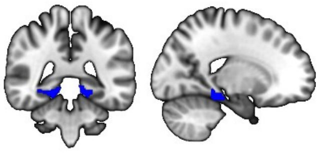 |
| BN_PM_PCun_dmPOS | Precuneus - dorsomedial parietooccipital sulcus | 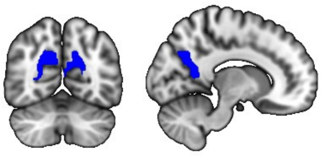 |
| BN_PM_CG_A23v | Cingulate gyrus - ventral area 23 | 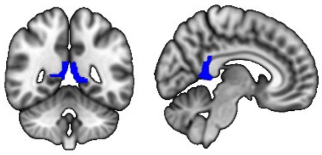 |
| BN_PM_OrG_A14m | Orbital gyrus - medial area 14 | 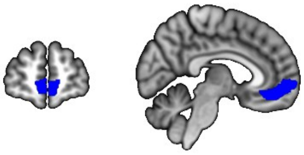 |

| | | |
|---|---|---|
| BN_PM_PCun_A31 | Precuneus - area 31 | 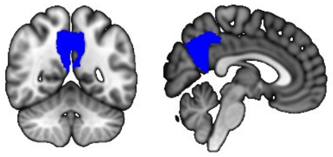 |
| BN_PM_SFG_A8dl | Superior frontal gyrus - dorsolateral area 8 | 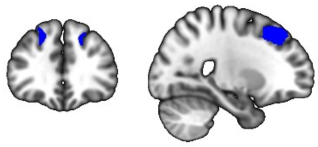 |
| BN_PM_MFG_A9_46d | Middle frontal gyrus - dorsal area 9/46 | 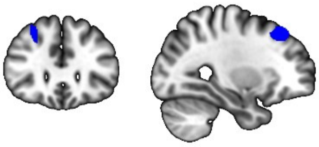 |
| BN_PM_MFG_A8vl | Middle frontal gyrus - ventrolateral area 8 | 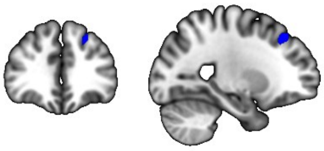 |
| BN_PM_IPL_A39rv | Inferior parietal lobule - rostroventral area 39 | 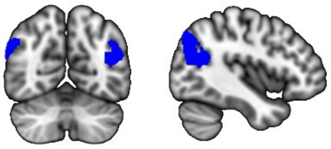 |
| BN_PM_IPL_A39rd | Inferior parietal lobule - rostrodorsal area 39 | 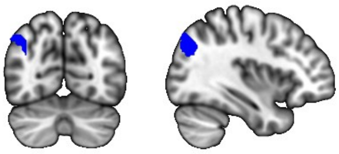 |
| BN_PM_OrG_A11m | Orbital gyrus – medial area 11 | 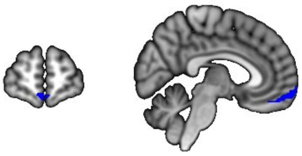 |
| BN_PM_CG_A23c | Cingulate gyrus - caudal area 23 | 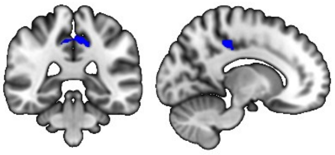 |
| BN_PM_IPL_A39c | Inferior parietal lobule - caudal area 39 | 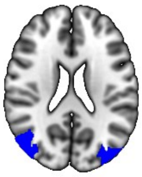 |
| BN_PM_CG_A23d | Cingulate gyrus - dorsal area 23 | 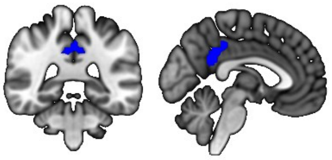 |
| BN_PM_Hipp_rHipp | Hippocampus – rostral hippocampus | 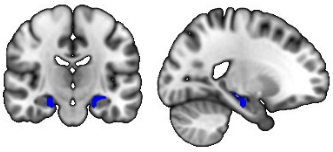 |

| | | |
|---|---|---|
| BN_PM_PhG_A28_34 | Parahippocampal gyrus - area 28/34 (entorhinal cortex) | 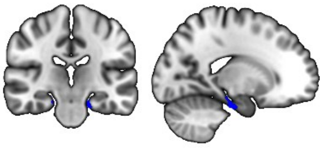 |
| BN_PM_OrG_A13 | Orbital gyrus - area 13 | 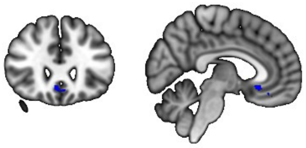 |
| BN_PM_PCun_A7m | Precuneus - medial area 7 | 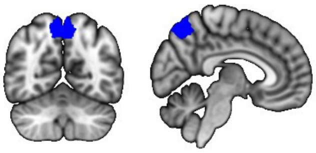 |
| BN_PM_PhG_A35_36c | Parahippocampal gyrus - caudal area 35/36 | 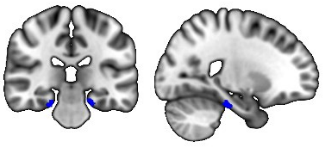 |
| BN_PM_SFG_A10m | Superior frontal gyrus - medial area 10 | 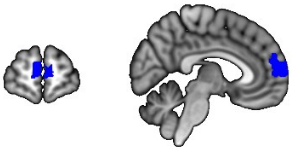 |
| BN_PM_MVOcC_rLinG | Medio ventral occipital cortex - rostral lingual gyrus | 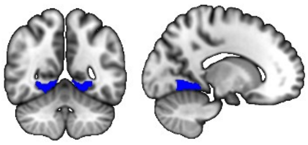 |
| BN_PM_MVOcC_vmPOS | Medio ventral occipital cortex - ventromedial parietooccipital sulcus | 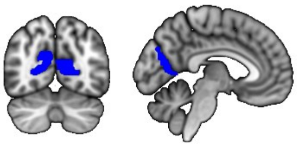 |
| BN_PM_MFG_A6vl | Middle frontal gyrus - ventrolateral area 6 | 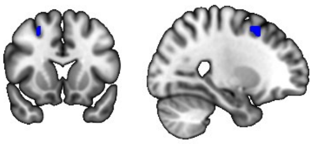 |
| BN_PM_PCun_A5m | Precuneus - medial area 5 | 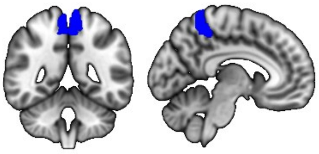 |
| BN_PM_MTG_A37dl | Middle temporal gyrus - dorsolateral area 37 | 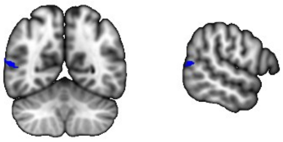 |
| BN_PM_SPL_A7r | Superior parietal lobule - rostral area 7 | 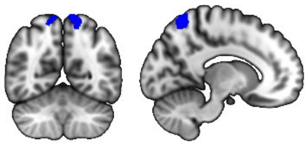 |

| | | |
|---|---|---|
| BN_PM_CG_A32sg | Cingulate gyrus - subgenual area 32 | 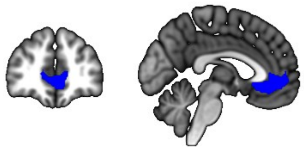 |
| BN_PM_CG_A24rv | Cingulate gyrus - rostroventral area 24 | 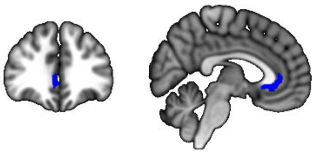 |
| BN_PM_FuG_A37mv | Fusiform gyrus - medioventral area 37 | 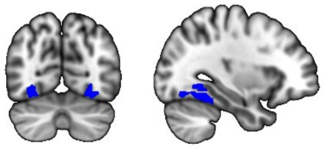 |
| BN_PM_Tha_Otha | Thalamus – occipital thalamus | 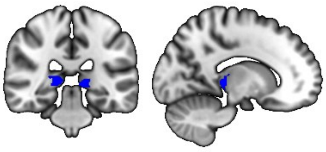 |
| BN_PM_SFG_A9m | Superior frontal gyrus - medial area 9 | 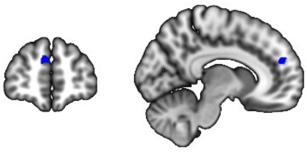 |
| BN_PM_LOcC_lsOccG | Lateral occipital cortex - lateral superior occipital gyrus | 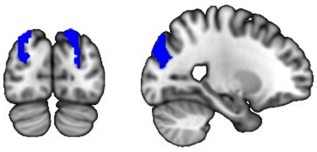 |
| BN_PM_Tha_cTtha | Thalamus - caudal temporal thalamus | 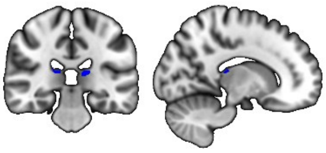 |
| BN_PM_Hipp_cHipp | Hippocampus - caudal hippocampus | 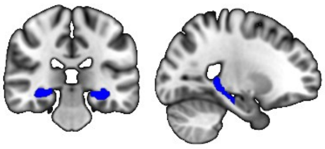 |
| BN_PM_LOcC_msOccG | Lateral occipital cortex - medial superior occipital gyrus | 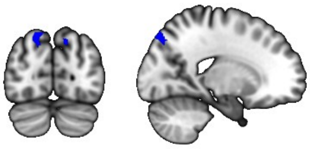 |
| BN_PM_Tha_PPtha | Thalamus - posterior parietal thalamus | 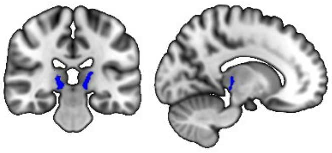 |
| BN_PM_FuG_A37lv | Fusiform gyrus - lateroventral area 37 | 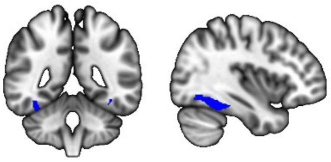 |

| | | |
|---|---|---|
| BN_PM_SPL_A7c | Superior parietal lobule - caudal area 7 | 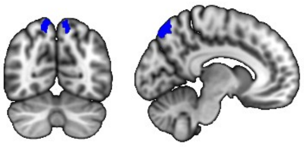 |
| BN_PM_PCL_A1_2_3ll | Paracentral lobule – area 1/2/3 (lower limb region) | 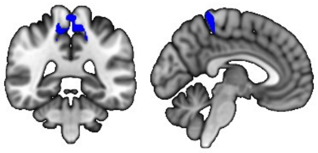 |
| BN_PM_MVOcC_rCunG | Medioventral occipital cortex - rostral cuneus gyrus | 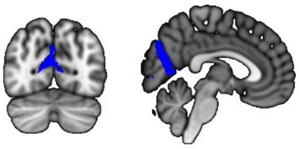 |
| BN_PM_LOcC_mOccG | Lateral occipital cortex – middle occipital gyrus | 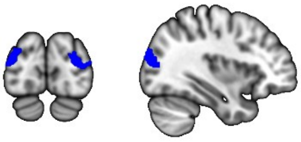 |
| BN_PM_LOcC_V5_MT | Lateral occipital cortex - area V5/MT+ | 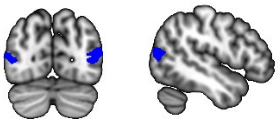 |
| BN_PM_LOcC_OPC | Lateral occipital cortex - occipital polar cortex | 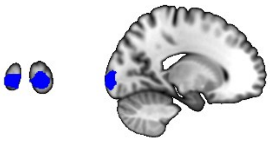 |
| BN_PM_MVOcC_cLinG | Medioventral occipital cortex - caudal lingual gyrus | 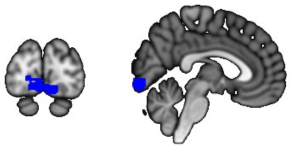 |
| BN_PM_MVOcC_cCunG | Medioventral occipital cortex - caudal cuneus gyrus | 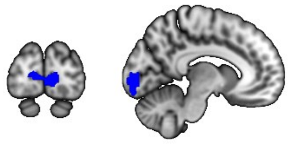 |
| BN_PM_LOcC_iOccG | Lateral occipital cortex - inferior occipital gyrus | 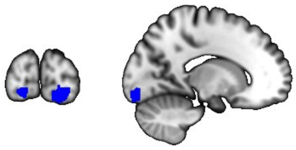 |

**Supplementary Table 3. Age-related functional changes over the adult lifespan in each region of interest (ROI) within the anterior-temporal (AT) network.** Statistical summary of the age term from linear mixed models adjusted for sex and education as F-test result. Trend was calculated as the trend of the estimated marginal means for functional changes in relation to age. Bolded *p*-values survived the Holm-Bonferroni correction ($p < 0.05$). Ddf: approximate denominator degrees of freedom.

| | ddf | *F* | trend | *P* | adjusted *P* |
|---|---|---|---|---|---|
| BN_AT_IPL_A39c | 189.51 | 34.54 | -0.0015 | 0.000 | **0.000** |
| BN_AT_LOcC_lsOccG | 192.20 | 34.07 | -0.0014 | 0.000 | **0.000** |
| BN_AT_IFG_IFS | 199.44 | 25.08 | -0.0013 | 0.000 | **0.000** |

| | | | | | |
|---|---|---|---|---|---|
| BN_AT_IPL_A39rd | 193.93 | 34.79 | -0.0012 | 0.000 | **0.000** |
| BN_AT_SPL_A7ip | 192.70 | 26.60 | -0.0012 | 0.000 | **0.000** |
| BN_AT_SPL_A7c | 190.57 | 25.26 | -0.0011 | 0.000 | **0.000** |
| BN_AT_ITG_A37vl | 197.43 | 17.93 | -0.0009 | 0.000 | **0.001** |
| BN_AT_ITG_A37elv | 193.59 | 15.69 | -0.0009 | 0.000 | **0.003** |
| BN_AT_FuG_A37lv | 190.76 | 14.90 | -0.0008 | 0.000 | **0.004** |
| BN_AT_MTG_A37dl | 178.00 | 14.69 | -0.0007 | 0.000 | **0.004** |
| BN_AT_PhG_TL | 199.93 | 3.13 | -0.0006 | 0.078 | 1.000 |
| BN_AT_OrG_A12_47o | 191.04 | 7.15 | -0.0006 | 0.008 | 0.171 |
| BN_AT_ITG_A20cl | 184.67 | 8.40 | -0.0005 | 0.004 | 0.092 |
| BN_AT_OrG_A11l | 204.65 | 2.77 | -0.0003 | 0.097 | 1.000 |
| BN_AT_LOcC_V5_MT | 191.48 | 1.85 | -0.0003 | 0.175 | 1.000 |
| BN_AT_Tha_rTtha | 188.44 | 2.40 | -0.0002 | 0.123 | 1.000 |
| BN_AT_MTG_A21c | 195.70 | 0.56 | -0.0002 | 0.456 | 1.000 |
| BN_AT_Amyg_mAmyg | 178.74 | 0.08 | -0.0001 | 0.783 | 1.000 |
| BN_AT_OrG_A13 | 196.52 | 0.04 | 0.0000 | 0.841 | 1.000 |
| BN_AT_STG_A38l | 200.78 | 0.00 | 0.0000 | 0.954 | 1.000 |
| BN_AT_Tha_mPFtha | 195.02 | 0.35 | 0.0001 | 0.554 | 1.000 |
| BN_AT_STG_A22r | 200.10 | 0.65 | 0.0001 | 0.421 | 1.000 |
| BN_AT_PhG_A35_36c | 206.45 | 0.23 | 0.0002 | 0.630 | 1.000 |
| BN_AT_BG_NAC | 182.50 | 2.09 | 0.0002 | 0.150 | 1.000 |
| BN_AT_ITG_A20cv | 211.94 | 0.91 | 0.0002 | 0.342 | 1.000 |
| BN_AT_BG_dlPu | 182.52 | 1.79 | 0.0002 | 0.183 | 1.000 |
| BN_AT_MTG_A21r | 197.52 | 1.42 | 0.0002 | 0.234 | 1.000 |
| BN_AT_Amyg_lAmyg | 198.51 | 1.69 | 0.0003 | 0.195 | 1.000 |
| BN_AT_BG_vmPu | 186.96 | 3.59 | 0.0003 | 0.060 | 1.000 |
| BN_AT_STG_TE10_TE12 | 181.50 | 4.27 | 0.0003 | 0.040 | 0.730 |
| BN_AT_INS_vId_vIg | 201.08 | 4.34 | 0.0003 | 0.038 | 0.730 |
| BN_AT_INS_vIa | 197.75 | 4.92 | 0.0005 | 0.028 | 0.553 |
| BN_AT_ITG_A20il | 204.02 | 9.50 | 0.0007 | 0.002 | 0.054 |
| BN_AT_Hipp_rHipp | 199.23 | 11.49 | 0.0008 | 0.001 | **0.020** |
| BN_AT_Hipp_cHipp | 193.49 | 15.44 | 0.0010 | 0.000 | **0.003** |
| BN_AT_ITG_A20iv | 210.15 | 14.95 | 0.0011 | 0.000 | **0.004** |
| BN_AT_ITG_A20r | 200.09 | 25.78 | 0.0011 | 0.000 | **0.000** |
| BN_AT_STG_A38m | 207.35 | 22.29 | 0.0012 | 0.000 | **0.000** |
| BN_AT_FuG_A20rv | 205.74 | 18.39 | 0.0012 | 0.000 | **0.001** |
| BN_AT_PhG_A35_36r | 203.47 | 16.82 | 0.0013 | 0.000 | **0.002** |
| BN_AT_PhG_TI | 205.42 | 27.93 | 0.0015 | 0.000 | **0.000** |
| BN_AT_PhG_A28_34 | 204.90 | 22.15 | 0.0016 | 0.000 | **0.000** |

**Supplementary Table 4. Age-related functional changes over the adult lifespan in each region of interest (ROI) within the posterior-medial (PM) network.** Statistical summary of the age term from linear mixed models adjusted for sex and education as F-test result. Trend was calculated as the trend of the estimated marginal means for functional changes in relation to age. Bolded *p*-values survived the Holm-Bonferroni correction ($p < 0.05$). Ddf: approximate denominator degrees of freedom.

| | ddf | *F* | trend | *P* | adjusted *P* |
|---|---|---|---|---|---|
| BN_PM_PhG_TL | 197,52 | 43,85 | -0,0023 | 0,000 | **0,000** |
| BN_PM_FuG_A20rv | 199,74 | 37,71 | -0,0022 | 0,000 | **0,000** |
| BN_PM_PhG_TH | 206,85 | 35,96 | -0,0021 | 0,000 | **0,000** |
| BN_PM_PCun_dmPOS | 207,55 | 35,24 | -0,0020 | 0,000 | **0,000** |
| BN_PM_CG_A23v | 199,35 | 34,20 | -0,0017 | 0,000 | **0,000** |
| BN_PM_OrG_A14m | 199,43 | 27,10 | -0,0014 | 0,000 | **0,000** |
| BN_PM_PCun_A31 | 204,50 | 28,71 | -0,0014 | 0,000 | **0,000** |
| BN_PM_SFG_A8dl | 198,94 | 23,02 | -0,0013 | 0,000 | **0,000** |
| BN_PM_MFG_A9_46d | 198,43 | 18,47 | -0,0013 | 0,000 | **0,001** |
| BN_PM_MFG_A8vl | 206,53 | 11,44 | -0,0012 | 0,001 | **0,026** |
| BN_PM_IPL_A39rv | 201,84 | 24,77 | -0,0012 | 0,000 | **0,000** |
| BN_PM_IPL_A39rd | 199,95 | 10,13 | -0,0012 | 0,002 | **0,049** |
| BN_PM_OrG_A11m | 198,99 | 18,40 | -0,0011 | 0,000 | **0,001** |
| BN_PM_CG_A23c | 187,04 | 26,88 | -0,0011 | 0,000 | **0,000** |
| BN_PM_IPL_A39c | 202,16 | 17,77 | -0,0011 | 0,000 | **0,001** |
| BN_PM_CG_A23d | 204,90 | 16,35 | -0,0011 | 0,000 | **0,003** |
| BN_PM_Hipp_rHipp | 209,23 | 12,01 | -0,0010 | 0,001 | **0,020** |
| BN_PM_PhG_A28_34 | 192,73 | 12,23 | -0,0010 | 0,001 | **0,019** |
| BN_PM_OrG_A13 | 196,38 | 17,38 | -0,0009 | 0,000 | **0,002** |
| BN_PM_PCun_A7m | 197,39 | 9,98 | -0,0009 | 0,002 | 0,051 |
| BN_PM_PhG_A35_36c | 201,77 | 9,97 | -0,0008 | 0,002 | 0,051 |
| BN_PM_SFG_A10m | 200,92 | 12,71 | -0,0008 | 0,000 | **0,015** |
| BN_PM_MVOcC_rLinG | 202,49 | 6,12 | -0,0007 | 0,014 | 0,269 |
| BN_PM_MVOcC_vmPOS | 205,51 | 7,75 | -0,0007 | 0,006 | 0,135 |
| BN_PM_MFG_A6vl | 194,29 | 2,90 | -0,0006 | 0,090 | 0,972 |
| BN_PM_PCun_A5m | 196,67 | 8,30 | -0,0006 | 0,004 | 0,106 |
| BN_PM_MTG_A37dl | 205,16 | 3,12 | -0,0006 | 0,079 | 0,947 |
| BN_PM_SPL_A7r | 186,68 | 5,47 | -0,0005 | 0,020 | 0,327 |
| BN_PM_CG_A32sg | 197,91 | 7,21 | -0,0005 | 0,008 | 0,159 |
| BN_PM_CG_A24rv | 194,46 | 9,45 | -0,0005 | 0,002 | 0,063 |
| BN_PM_FuG_A37mv | 207,16 | 2,93 | -0,0005 | 0,088 | 0,972 |
| BN_PM_Tha_Otha | 206,26 | 8,54 | -0,0004 | 0,004 | 0,096 |
| BN_PM_SFG_A9m | 212,47 | 2,75 | -0,0004 | 0,099 | 0,972 |
| BN_PM_LOcC_lsOccG | 198,96 | 2,86 | -0,0004 | 0,093 | 0,972 |
| BN_PM_Tha_cTtha | 201,25 | 7,28 | -0,0004 | 0,008 | 0,159 |
| BN_PM_Hipp_cHipp | 205,14 | 4,59 | -0,0004 | 0,033 | 0,466 |
| BN_PM_LOcC_msOccG | 187,37 | 2,05 | -0,0004 | 0,154 | 1,000 |
| BN_PM_Tha_PPtha | 197,02 | 5,88 | -0,0003 | 0,016 | 0,291 |
| BN_PM_FuG_A37lv | 204,24 | 1,39 | -0,0003 | 0,240 | 1,000 |
| BN_PM_SPL_A7c | 195,84 | 1,06 | -0,0003 | 0,304 | 1,000 |
| BN_PM_PCL_A1_2_3ll | 196,60 | 0,11 | -0,0001 | 0,746 | 1,000 |

| | | | | | |
|---|---|---|---|---|---|
| BN_PM_MVOcC_rCunG | 189,41 | 0,27 | 0,0001 | 0,607 | 1,000 |
| BN_PM_LOcC_mOccG | 198,82 | 0,47 | 0,0002 | 0,493 | 1,000 |
| BN_PM_LOcC_V5_MT | 200,03 | 0,57 | 0,0002 | 0,453 | 1,000 |
| BN_PM_LOcC_OPC | 180,70 | 4,28 | 0,0004 | 0,040 | 0,521 |
| BN_PM_MVOcC_cLinG | 191,68 | 5,44 | 0,0005 | 0,021 | 0,327 |
| BN_PM_MVOcC_cCunG | 177,66 | 5,85 | 0,0005 | 0,017 | 0,291 |
| BN_PM_LOcC_iOccG | 192,44 | 7,73 | 0,0006 | 0,006 | 0,135 |

**<u>Supplementary Table 1.</u> Summary of Brainnetome-based regions of interest (ROIs) included in the anterior-temporal (AT) mask derived from Aβ-negative cognitively unimpaired adults over the age of 60.**

| ROI | Brainnetome labeling | Brain view |
|---|---|---|
| BN_AT_ITG_A20il | Inferior temporal gyrus - intermediate lateral area 20 | 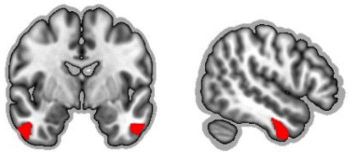 |
| BN_AT_ITG_A20iv | Inferior temporal gyrus - intermediate ventral area 20 | 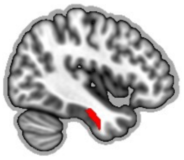 |
| BN_AT_ITG_A20r | Inferior temporal gyrus - rostral area 20 | 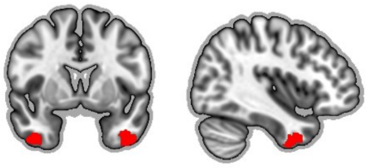 |
| BN_AT_STG_TE10_TE12 | Superior temporal gyrus - TE1.0 and TE1.2 | 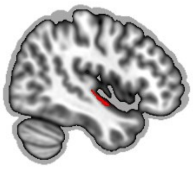 |
| BN_AT_MTG_A21r | Middle temporal gyrus - rostral area 21 | 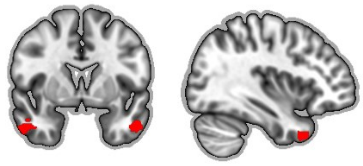 |

| | | |
|---|---|---|
| BN_AT_Hipp_cHipp | Caudal hippocampus | 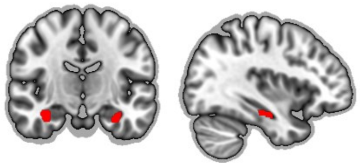 |
| BN_AT_FuG_A20rv | Fusiform gyrus - rostroventral area 20 | 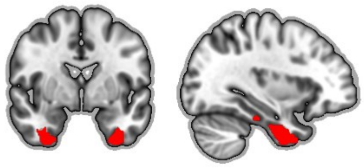 |
| BN_AT_PhG_A28_34 | Parahippocampal gyrus - entorhinal cortex | 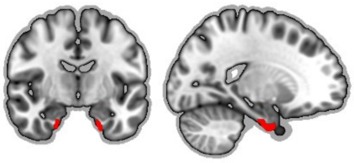 |
| BN_AT_PhG_TL | Parahippocampal gyrus – lateral posterior parahippocampal gyrus | 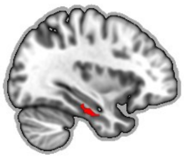 |
| BN_AT_PhG_A35_36r | Parahippocampal gyrus - rostral area 35/36 | 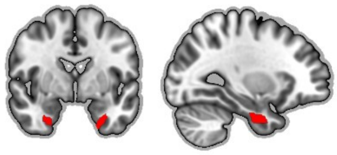 |
| BN_AT_Hipp_rHipp | Rostral hippocampus | 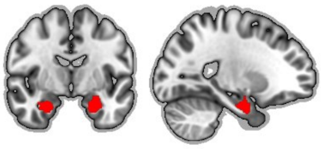 |
| BN_AT_STG_A38l | Superior temporal gyrus - lateral area 38 | 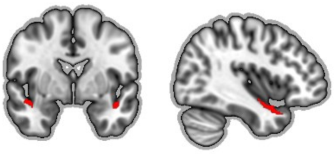 |
| BN_AT_INS_vId_vIg | Ventral dysgranular and granular insula | 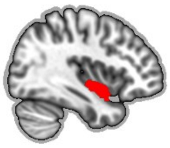 |

| | | |
|---|---|---|
| BN_AT_PhG_TI | Parahippocampal gyrus - temporal agranular insular cortex | 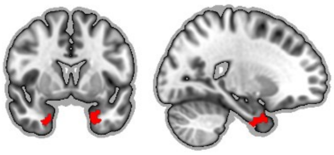 |
| BN_AT_STG_A38m | Superior temporal gyrus - medial area 38 | 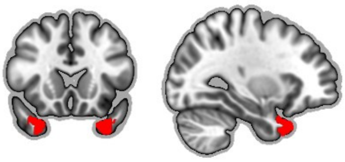 |
| BN_AT_Amyg_mAmyg | Medial amygdala | 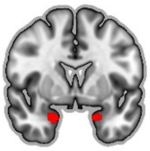 |
| BN_AT_PhG_A35_36c | Parahippocampal gyrus - caudal area 35/36 | 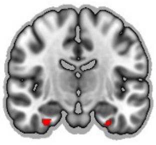 |
| BN_AT_Amyg_lAmyg | Lateral amygdala | 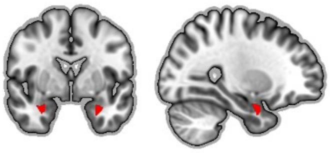 |
| BN_AT_BG_NAC | Nucleus accumbens | 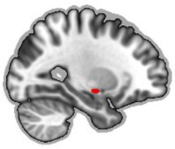 |
| BN_AT_OrG_A12_47o | Orbital gyrus - orbital area 12/47 | 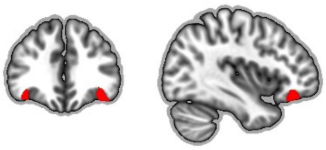 |
| BN_AT_ITG_A37elv | Inferior temporal gyrus - extreme lateroventral area 37 | 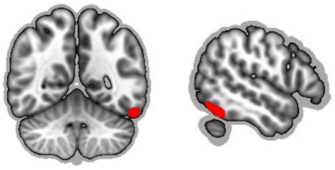 |

| | | |
|---|---|---|
| BN_AT_OrG_A11l | Orbital gyrus - lateral area 11 | 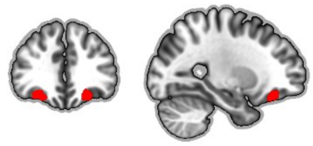 |
| BN_AT_ITG_A37vl | Inferior temporal gyrus - ventrolateral area 37 | 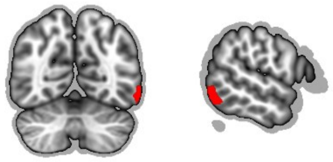 |
| BN_AT_FuG_A37lv | Fusiform gyrus - lateroventral area 37 | 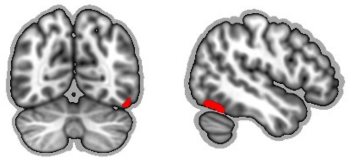 |

**<u>Supplementary Table 2.</u> Summary of Brainnetome-based regions of interest (ROIs) included in the posterior-medial (PM) mask derived from Aβ-negative cognitively unimpaired adults over the age of 60.**

| ROI | Brainnetome labeling | Brain view |
|---|---|---|
| BN_PM_PhG_A35_36c | Parahippocampal gyrus - caudal area 35/36 | 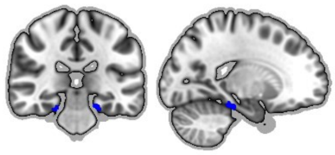 |
| BN_PM_FuG_A37mv | Fusiform gyrus - medioventral area37 | 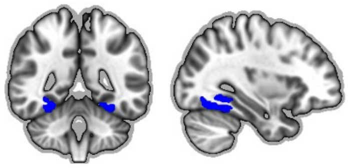 |
| BN_PM_MVOcC_rLinG | Medio ventral occipital cortex - rostral lingual gyrus | 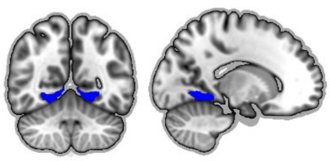 |
| BN_PM_LOcC_iOccG | Lateral occipital cortex - inferior occipital gyrus | 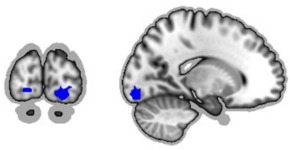 |

| | | |
|---|---|---|
| BN_PM_Tha_Otha | Occipital thalamus | 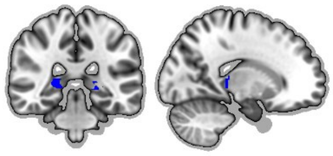 |
| BN_PM_PhG_TH | Parahippocampal gyrus - medial posterior parahippocampal gyrus | 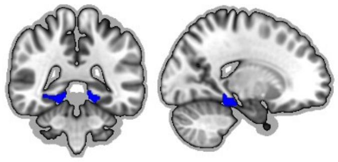 |
| BN_PM_FuG_A37lv | Fusiform gyrus - lateroventral area 37 | 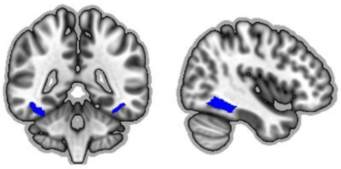 |
| BN_PM_MVOcC_cLinG | Medio-ventral occipital cortex - caudal lingual gyrus | 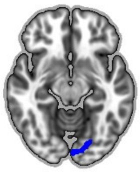 |
| BN_PM_Hipp_cHipp | Caudal hippocampus | 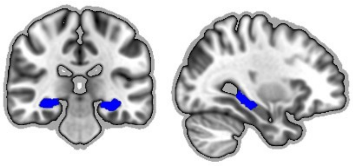 |
| BN_PM_LOcC_OPC | Lateral occipital cortex - occipital polar cortex | 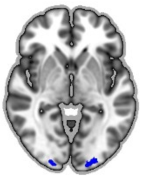 |
| BN_PM_FuG_A20rv | Fusiform gyrus - rostroventral area 20 | 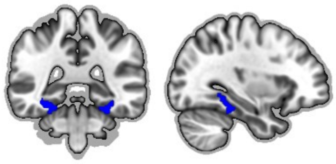 |
| BN_PM_MVOcC_cCunG | Medio-ventral occipital cortex - caudal cuneus gyrus | 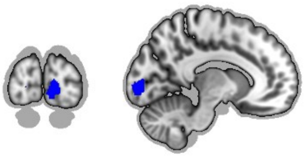 |

| | | |
|---|---|---|
| BN_PM_LOcC_mOccG | Lateral occipital cortex - medial superior occipital gyrus | 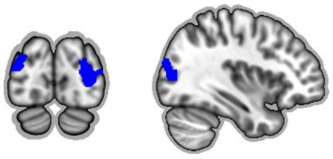 |
| BN_PM_PhG_TL | Parahippocampal gyrus - lateral posterior parahippocampal gyrus | 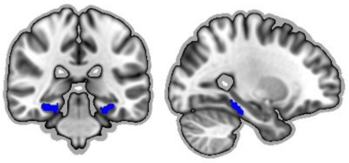 |
| BN_PM_ITG_A37vl | Inferior temporal gyrus - ventrolateral area 37 | 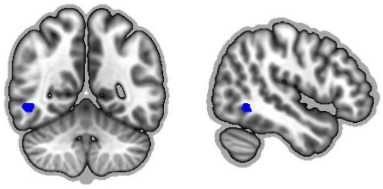 |
| BN_PM_Hipp_rHipp | Rostral hippocampus | 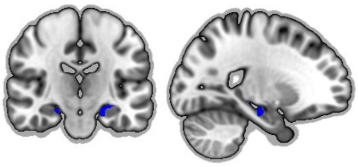 |
| BN_PM_LOcC_V5_MT | Lateral occipital cortex - area V5/MT+ | 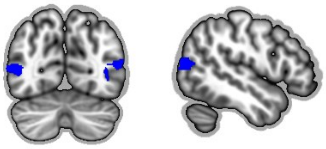 |
| BN_PM_PCL_A1_2_3ll | Paracentral lobule - area1/2/3 (lower limb region) | 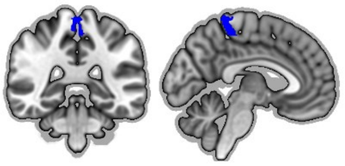 |
| BN_PM_OrG_A11m | Orbital gyrus - medial area 11 | 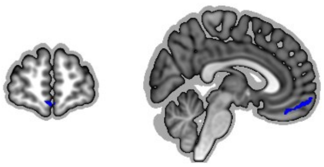 |
| BN_PM_MVOcC_rCunG | Medio-ventral occipital cortex - rostral cuneus gyrus | 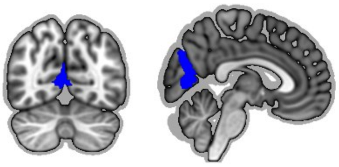 |
| BN_PM_PCL_A4ll | Paracentral lobule - area 4 (lower limb region) | 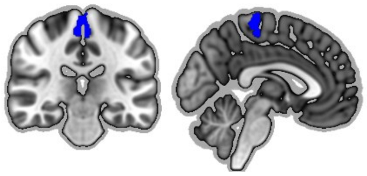 |

| | | |
|---|---|---|
| BN_PM_CG_A23v | Cingulate gyrus - ventral area 23 | 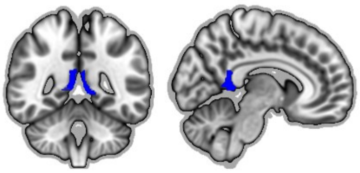 |
| BN_PM_CG_A32sg | Cingulate gyrus - subgenual area 32 | 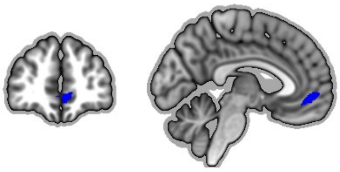 |
| BN_PM_PCun_A31 | Precuneus - area 31 (Lc1) | 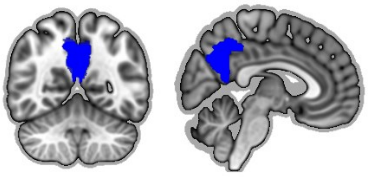 |
| BN_PM_LOcC_lsOccG | Lateral occipital cortex - lateral superior occipital gyrus | 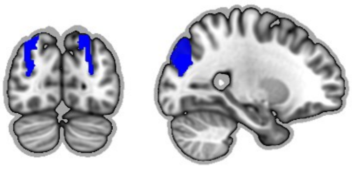 |
| BN_PM_PCun_A5m | Precuneus - medial area 5(PEm) | 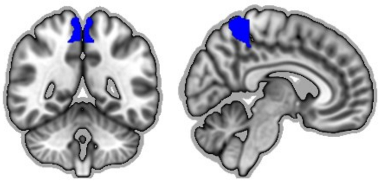 |
| BN_PM_LOcC_msOccG | Lateral occipital cortex - medial superior occipital gyrus | 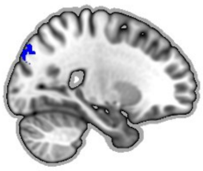 |
| BN_PM_OrG_A14m | Orbital gyrus - medial area 14 | 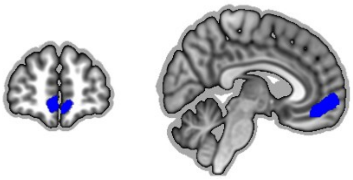 |
| BN_PM_CG_A23d | Cingulate gyrus - dorsal area 23 | 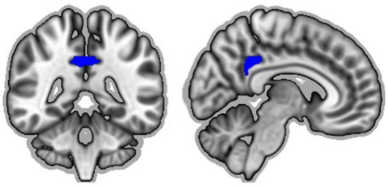 |
| BN_PM_IPL_A39c | Inferior parietal lobule - caudal area 39 (PGp) | 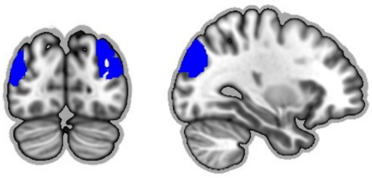 |

| | | |
|---|---|---|
| BN_PM_MVOcC_vmPOS | Medio-ventral occipital cortex - ventromedial parietooccipital sulcus | 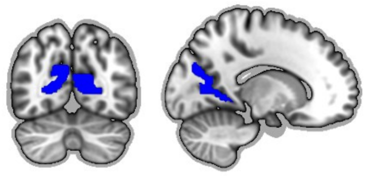 |
| BN_PM_SPL_A7r | Superior parietal lobule - rostral area 7 | 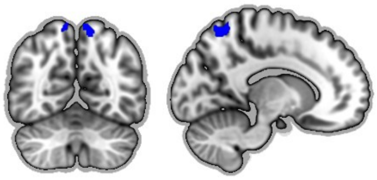 |
| BN_PM_MFG_A6vl | Middle frontal gyrus - ventrolateral area 6 | 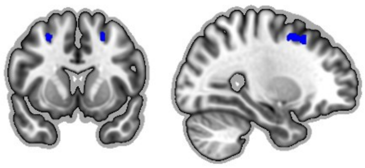 |
| BN_PM_IPL_A39rv | Inferior parietal lobule - rostroventral area 39 (PGa) | 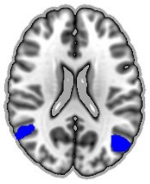 |
| BN_PM_MFG_A9_46d | Middle frontal gyrus - dorsal area 9/46 | 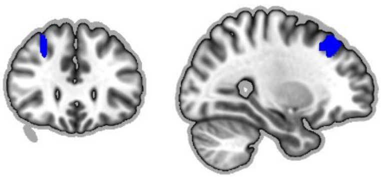 |
| BN_PM_PCun_A7m | Precuneus - medial area 7 (PEp) | 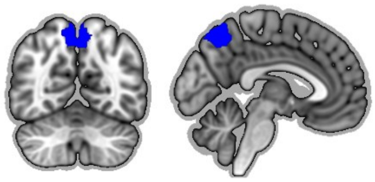 |
| BN_PM_PCun_dmPOS | Precuneus - dorsomedial parietooccipital sulcus (PEr) | 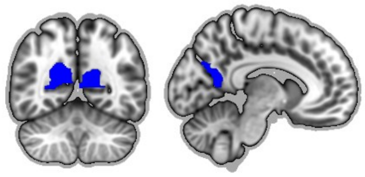 |
| BN_PM_MFG_A8vl | Middle frontal gyrus - ventrolateral area 8 | 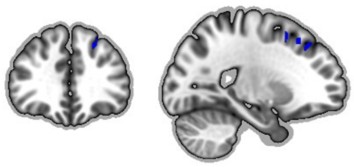 |
| BN_PM_IPL_A39rd | Inferior parietal lobule - rostrodorsal area 39 (Hip3) | 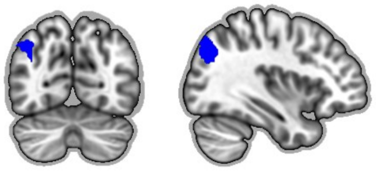 |

| | | |
|---|---|---|
| BN_PM_SFG_A8dl | Superior frontal gyrus - dorsolateral area 8 | 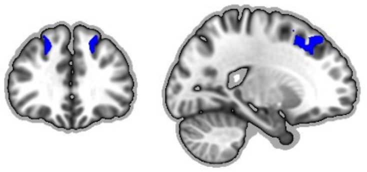 |
| BN_PM_SPL_A7c | Superior parietal lobule - caudal area 7 | 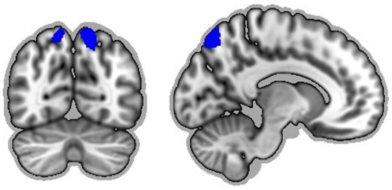 |

**Supplementary Table 3. Functional changes related to amyloid uptake in each region of interest (ROI) within the anterior-temporal (AT) network.** Statistical summary of amyloid uptake term from linear mixed models adjusted for age, sex, education and time as F-test result. Trend was calculated as the trend of the estimated marginal means for functional changes in relation to amyloid burden. Bolded *p*-values survived the Holm-Bonferroni correction ($p < 0.05$). Ddf: approximate denominator degrees of freedom.

| | ddf | *F* | trend | *P* | adjusted *P* |
|---|---|---|---|---|---|
| BN_AT_ITG_A20il | 130.15 | 24.61 | 0.2455 | 0.000 | **0.000** |
| BN_AT_ITG_A20iv | 128.76 | 20.32 | 0.3025 | 0.000 | **0.000** |
| BN_AT_ITG_A20r | 127.62 | 16.99 | 0.1907 | 0.000 | **0.001** |
| BN_AT_STG_TE10_TE12 | 200.00 | 16.61 | 0.1390 | 0.000 | **0.001** |
| BN_AT_MTG_A21r | 129.92 | 15.29 | 0.1847 | 0.000 | **0.003** |
| BN_AT_Hipp_cHipp | 133.03 | 13.22 | 0.2295 | 0.000 | **0.007** |
| BN_AT_FuG_A20rv | 131.96 | 12.34 | 0.2052 | 0.001 | **0.011** |
| BN_AT_PhG_A28_34 | 135.20 | 11.27 | 0.2533 | 0.001 | **0.017** |
| BN_AT_PhG_TL | 127.52 | 9.87 | 0.1943 | 0.002 | **0.033** |
| BN_AT_PhG_A35_36r | 131.07 | 9.87 | 0.2131 | 0.002 | **0.033** |
| BN_AT_Hipp_rHipp | 133.17 | 8.84 | 0.1673 | 0.004 | **0.049** |
| BN_AT_STG_A38l | 131.15 | 7.61 | 0.1001 | 0.007 | 0.086 |
| BN_AT_INS_vld_vlg | 135.66 | 7.56 | 0.0851 | 0.007 | 0.086 |
| BN_AT_PhG_TI | 134.29 | 5.83 | 0.1555 | 0.017 | 0.188 |
| BN_AT_STG_A38m | 133.24 | 5.47 | 0.1147 | 0.021 | 0.209 |
| BN_AT_Amyg_mAmyg | 128.82 | 5.03 | 0.1134 | 0.027 | 0.239 |
| BN_AT_PhG_A35_36c | 129.79 | 4.76 | 0.1622 | 0.031 | 0.248 |
| BN_AT_Amyg_lAmyg | 130.88 | 4.59 | 0.1107 | 0.034 | 0.248 |
| BN_AT_BG_NAC | 200.00 | 1.64 | 0.0419 | 0.202 | 1.000 |
| BN_AT_OrG_A12_47o | 131.99 | 1.53 | 0.0485 | 0.218 | 1.000 |
| BN_AT_ITG_A37elv | 132.26 | 0.18 | -0.0192 | 0.669 | 1.000 |
| BN_AT_OrG_A11l | 129.23 | 0.15 | 0.0150 | 0.695 | 1.000 |
| BN_AT_ITG_A37vl | 134.69 | 0.13 | -0.0166 | 0.714 | 1.000 |
| BN_AT_FuG_A37lv | 127.41 | 0.01 | -0.0042 | 0.911 | 1.000 |

**<u>Supplementary Table 4.</u> Functional changes related to glucose metabolism in each region of interest (ROI) within the anterior-temporal (AT) network.** Statistical summary of glucose metabolism term from linear mixed models adjusted for age, sex, education and time as F-test result. Trend was calculated as the trend of the estimated marginal means for functional changes in relation to glucose metabolism. Bolded *p*-values survived the Holm-Bonferroni correction ($p < 0.05$). Ddf: approximate denominator degrees of freedom.

| | ddf | *F* | trend | *P* | adjusted *P* |
|---|---|---|---|---|---|
| BN_AT_ITG_A20il | 126.67 | 31.42 | -0.1429 | 0.000 | **0.000** |
| BN_AT_Hipp_cHipp | 132.05 | 29.30 | -0.1663 | 0.000 | **0.000** |
| BN_AT_MTG_A21r | 125.62 | 28.14 | -0.1212 | 0.000 | **0.000** |
| BN_AT_Hipp_rHipp | 130.00 | 22.07 | -0.1291 | 0.000 | **0.000** |
| BN_AT_STG_TE10_TE12 | 212.00 | 21.17 | -0.0786 | 0.000 | **0.000** |
| BN_AT_ITG_A20r | 123.06 | 20.94 | -0.1080 | 0.000 | **0.000** |
| BN_AT_Amyg_lAmyg | 125.71 | 18.09 | -0.1061 | 0.000 | **0.001** |
| BN_AT_PhG_A35_36r | 128.64 | 18.08 | -0.1439 | 0.000 | **0.001** |
| BN_AT_ITG_A20iv | 130.89 | 15.22 | -0.1384 | 0.000 | **0.002** |
| BN_AT_Amyg_mAmyg | 128.84 | 14.32 | -0.0949 | 0.000 | **0.004** |
| BN_AT_PhG_A28_34 | 133.96 | 13.86 | -0.1409 | 0.000 | **0.004** |
| BN_AT_FuG_A20rv | 131.37 | 13.83 | -0.1116 | 0.000 | **0.004** |
| BN_AT_INS_vId_vIg | 129.94 | 13.74 | -0.0574 | 0.000 | **0.004** |
| BN_AT_PhG_A35_36c | 130.65 | 12.91 | -0.1335 | 0.000 | **0.005** |
| BN_AT_STG_A38l | 131.28 | 11.49 | -0.0616 | 0.001 | **0.009** |
| BN_AT_PhG_TI | 133.64 | 10.05 | -0.1048 | 0.002 | **0.017** |
| BN_AT_PhG_TL | 132.35 | 10.01 | -0.0967 | 0.002 | **0.017** |
| BN_AT_STG_A38m | 131.58 | 8.47 | -0.0736 | 0.004 | **0.030** |
| BN_AT_BG_NAC | 212.00 | 6.43 | -0.0410 | 0.012 | 0.072 |
| BN_AT_OrG_A12_47o | 127.63 | 4.42 | -0.0408 | 0.037 | 0.187 |
| BN_AT_OrG_A11l | 118.87 | 4.17 | -0.0382 | 0.043 | 0.187 |
| BN_AT_FuG_A37lv | 124.08 | 1.61 | -0.0242 | 0.207 | 0.620 |
| BN_AT_ITG_A37elv | 131.65 | 0.37 | -0.0141 | 0.545 | 1.000 |
| BN_AT_ITG_A37vl | 127.79 | 0.02 | -0.0034 | 0.884 | 1.000 |

**<u>Supplementary Table 5.</u> Functional changes related to hippocampal volume in each region of interest (ROI) within the anterior-temporal (AT) network.** Statistical summary of hippocampal volume term from linear mixed models adjusted for age, sex, education and time as F-test result. Trend was calculated as the trend of the estimated marginal means for functional changes in relation to hippocampal volume. Bolded *p*-values survived the Holm-Bonferroni correction ($p < 0.05$). Ddf: approximate denominator degrees of freedom.

| | ddf | *F* | trend | *P* | adjusted *P* |
|---|---|---|---|---|---|

| | | | | | |
|---|---|---|---|---|---|
| BN_AT_Hipp_cHipp | 113.39 | 21.92 | -0.1473 | 0.000 | **0.000** |
| BN_AT_Amyg_lAmyg | 112.80 | 13.78 | -0.0916 | 0.000 | **0.007** |
| BN_AT_Amyg_mAmyg | 107.80 | 11.36 | -0.0827 | 0.001 | **0.023** |
| BN_AT_PhG_A28_34 | 118.28 | 10.67 | -0.1271 | 0.001 | **0.030** |
| BN_AT_Hipp_rHipp | 113.86 | 10.20 | -0.0922 | 0.002 | **0.036** |
| BN_AT_STG_A38m | 119.52 | 9.63 | -0.0779 | 0.002 | **0.045** |
| BN_AT_ITG_A20r | 109.43 | 9.53 | -0.0753 | 0.003 | **0.046** |
| BN_AT_ITG_A20il | 112.32 | 9.25 | -0.0841 | 0.003 | **0.050** |
| BN_AT_MTG_A21r | 109.51 | 7.23 | -0.0654 | 0.008 | 0.133 |
| BN_AT_PhG_A35_36r | 112.35 | 6.86 | -0.0923 | 0.010 | 0.150 |
| BN_AT_PhG_TL | 113.18 | 6.78 | -0.0819 | 0.010 | 0.150 |
| BN_AT_STG_TE10_TE12 | 104.84 | 6.53 | -0.0422 | 0.012 | 0.157 |
| BN_AT_PhG_TI | 119.84 | 6.14 | -0.0819 | 0.015 | 0.175 |
| BN_AT_STG_A38l | 116.32 | 5.36 | -0.0434 | 0.022 | 0.246 |
| BN_AT_FuG_A20rv | 113.84 | 5.13 | -0.0700 | 0.025 | 0.253 |
| BN_AT_PhG_A35_36c | 114.31 | 4.45 | -0.0822 | 0.037 | 0.334 |
| BN_AT_ITG_A20iv | 112.18 | 3.68 | -0.0710 | 0.057 | 0.460 |
| BN_AT_OrG_A11l | 115.91 | 2.13 | 0.0288 | 0.147 | 1.000 |
| BN_AT_INS_vId_vIg | 116.88 | 1.81 | -0.0211 | 0.181 | 1.000 |
| BN_AT_BG_NAC | 103.34 | 1.25 | -0.0178 | 0.266 | 1.000 |
| BN_AT_OrG_A12_47o | 122.12 | 0.63 | 0.0168 | 0.431 | 1.000 |
| BN_AT_ITG_A37vl | 116.40 | 0.31 | 0.0126 | 0.579 | 1.000 |
| BN_AT_ITG_A37elv | 117.39 | 0.26 | 0.0116 | 0.613 | 1.000 |
| BN_AT_FuG_A37lv | 109.80 | 0.03 | -0.0030 | 0.872 | 1.000 |

**Supplementary Table 6. Functional changes related to Mini-Mental Scale Examination score (MMSE) in each region of interest (ROI) within the anterior-temporal (AT) network.** Statistical summary of MMSE term from linear mixed models adjusted for age, sex, education and time as F-test result. Trend was calculated as the trend of the estimated marginal means for functional changes in relation to MMSE. Bolded *p*-values survived the Holm-Bonferroni correction ($p < 0.05$). Ddf: approximate denominator degrees of freedom.

| | ddf | *F* | trend | *P* | adjusted *P* |
|---|---|---|---|---|---|
| BN_AT_ITG_A20il | 164.49 | 28.08 | -0.0086 | 0.000 | **0.000** |
| BN_AT_MTG_A21r | 159.78 | 23.11 | -0.0071 | 0.000 | **0.000** |
| BN_AT_STG_TE10_TE12 | 231.00 | 16.80 | -0.0046 | 0.000 | **0.001** |
| BN_AT_Hipp_cHipp | 171.10 | 12.97 | -0.0072 | 0.000 | **0.009** |
| BN_AT_INS_vId_vIg | 160.82 | 12.37 | -0.0035 | 0.001 | **0.011** |
| BN_AT_ITG_A20r | 158.66 | 11.62 | -0.0053 | 0.001 | **0.016** |
| BN_AT_STG_A38l | 161.00 | 11.01 | -0.0039 | 0.001 | **0.020** |
| BN_AT_ITG_A20iv | 163.44 | 8.80 | -0.0068 | 0.003 | 0.059 |

| | | | | | |
|---|---|---|---|---|---|
| BN_AT_Hipp_rHipp | 167.77 | 8.35 | -0.0053 | 0.004 | 0.070 |
| BN_AT_PhG_TL | 167.26 | 7.70 | -0.0054 | 0.006 | 0.092 |
| BN_AT_Amyg_lAmyg | 162.07 | 7.24 | -0.0044 | 0.008 | 0.110 |
| BN_AT_PhG_A35_36r | 165.00 | 7.00 | -0.0058 | 0.009 | 0.116 |
| BN_AT_PhG_A28_34 | 171.31 | 5.25 | -0.0056 | 0.023 | 0.278 |
| BN_AT_PhG_A35_36c | 166.57 | 5.19 | -0.0055 | 0.024 | 0.278 |
| BN_AT_FuG_A20rv | 166.23 | 5.14 | -0.0044 | 0.025 | 0.278 |
| BN_AT_Amyg_mAmyg | 161.57 | 4.72 | -0.0035 | 0.031 | 0.281 |
| BN_AT_PhG_TI | 167.47 | 4.67 | -0.0045 | 0.032 | 0.281 |
| BN_AT_BG_NAC | 150.78 | 3.68 | -0.0020 | 0.057 | 0.398 |
| BN_AT_FuG_A37lv | 155.68 | 2.72 | -0.0020 | 0.101 | 0.607 |
| BN_AT_STG_A38m | 166.26 | 1.99 | -0.0024 | 0.161 | 0.804 |
| BN_AT_OrG_A12_47o | 164.47 | 1.11 | -0.0014 | 0.294 | 1.000 |
| BN_AT_ITG_A37vl | 161.64 | 0.89 | -0.0014 | 0.347 | 1.000 |
| BN_AT_ITG_A37elv | 162.16 | 0.67 | -0.0012 | 0.413 | 1.000 |
| BN_AT_OrG_A11l | 158.82 | 0.01 | -0.0001 | 0.914 | 1.000 |

**Supplementary Table 7. Functional changes related to Mattis Dementia Rating Scale score (MDRS) in each region of interest (ROI) within the anterior-temporal (AT) network.** Statistical summary of MDRS term from linear mixed models adjusted for age, sex, education and time as F-test result. Trend was calculated as the trend of the estimated marginal means for functional changes in relation to MDRS. Bolded *p*-values survived the Holm-Bonferroni correction ($p < 0.05$). Ddf: approximate denominator degrees of freedom.

| | ddf | *F* | trend | *P* | adjusted *P* |
|---|---|---|---|---|---|
| BN_AT_ITG_A20il | 162.93 | 27.79 | -0.0031 | 0.000 | **0.000** |
| BN_AT_STG_TE10_TE12 | 160.11 | 17.11 | -0.0018 | 0.000 | **0.001** |
| BN_AT_INS_vId_vIg | 161.89 | 16.81 | -0.0016 | 0.000 | **0.001** |
| BN_AT_MTG_A21r | 159.00 | 15.90 | -0.0022 | 0.000 | **0.002** |
| BN_AT_Hipp_cHipp | 171.43 | 13.14 | -0.0026 | 0.000 | **0.008** |
| BN_AT_ITG_A20r | 159.28 | 12.69 | -0.0020 | 0.000 | **0.009** |
| BN_AT_Hipp_rHipp | 169.42 | 11.88 | -0.0023 | 0.001 | **0.013** |
| BN_AT_PhG_A35_36r | 166.62 | 9.92 | -0.0025 | 0.002 | **0.033** |
| BN_AT_PhG_A28_34 | 171.82 | 9.16 | -0.0027 | 0.003 | **0.046** |
| BN_AT_Amyg_lAmyg | 164.36 | 8.09 | -0.0017 | 0.005 | 0.075 |
| BN_AT_BG_NAC | 158.04 | 8.08 | -0.0011 | 0.005 | 0.075 |
| BN_AT_PhG_TL | 169.49 | 6.92 | -0.0019 | 0.009 | 0.121 |
| BN_AT_ITG_A20iv | 160.26 | 6.59 | -0.0022 | 0.011 | 0.134 |
| BN_AT_FuG_A20rv | 166.83 | 6.43 | -0.0018 | 0.012 | 0.134 |
| BN_AT_STG_A38l | 158.51 | 6.22 | -0.0011 | 0.014 | 0.136 |
| BN_AT_PhG_TI | 169.08 | 6.00 | -0.0019 | 0.015 | 0.138 |

| | | | | | |
|---|---|---|---|---|---|
| BN_AT_Amyg_mAmyg | 165.22 | 5.50 | -0.0014 | 0.020 | 0.162 |
| BN_AT_PhG_A35_36c | 169.56 | 5.38 | -0.0021 | 0.022 | 0.162 |
| BN_AT_FuG_A37lv | 158.82 | 2.39 | -0.0007 | 0.124 | 0.746 |
| BN_AT_OrG_A12_47o | 168.22 | 1.90 | -0.0007 | 0.170 | 0.850 |
| BN_AT_STG_A38m | 168.02 | 1.73 | -0.0008 | 0.190 | 0.850 |
| BN_AT_ITG_A37vl | 166.52 | 1.11 | -0.0006 | 0.295 | 0.884 |
| BN_AT_ITG_A37elv | 166.78 | 1.02 | -0.0006 | 0.314 | 0.884 |
| BN_AT_OrG_A11l | 162.71 | 0.51 | -0.0004 | 0.475 | 0.884 |

**Supplementary Table 8. Functional changes related to amyloid uptake in each region of interest (ROI) within the posterior-medial (PM) network.** Statistical summary of amyloid uptake term from linear mixed models adjusted for age, sex, education and time as F-test result. Trend was calculated as the trend of the estimated marginal means for functional changes in relation to amyloid burden. Bolded *p*-values survived the Holm-Bonferroni correction ($p < 0.05$). Ddf: approximate denominator degrees of freedom.

| | ddf | *F* | trend | *P* | adjusted *P* |
|---|---|---|---|---|---|
| BN_PM_PhG_A35_36c | 134.23 | 20.04 | 0.2364 | 0.000 | **0.001** |
| BN_PM_FuG_A37mv | 138.89 | 18.47 | 0.2137 | 0.000 | **0.001** |
| BN_PM_MVOcC_rLinG | 133.48 | 15.13 | 0.2036 | 0.000 | **0.006** |
| BN_PM_Tha_Otha | 200.00 | 11.72 | 0.1556 | 0.001 | **0.029** |
| BN_PM_LOcC_iOccG | 129.60 | 11.67 | 0.1723 | 0.001 | **0.031** |
| BN_PM_FuG_A37lv | 132.15 | 10.96 | 0.1439 | 0.001 | **0.043** |
| BN_PM_SFG_A8dl | 124.55 | 10.63 | -0.1161 | 0.001 | 0.050 |
| BN_PM_PhG_TH | 134.90 | 9.34 | 0.1769 | 0.003 | 0.092 |
| BN_PM_IPL_A39rd | 120.49 | 7.69 | -0.1420 | 0.006 | 0.212 |
| BN_PM_SPL_A7c | 133.80 | 7.41 | -0.1125 | 0.007 | 0.235 |
| BN_PM_LOcC_mOccG | 125.04 | 6.42 | 0.1236 | 0.013 | 0.388 |
| BN_PM_MFG_A8vl | 125.18 | 6.40 | -0.1086 | 0.013 | 0.388 |
| BN_PM_MVOcC_cLinG | 125.87 | 5.81 | 0.1207 | 0.017 | 0.505 |
| BN_PM_FuG_A20rv | 136.13 | 5.68 | 0.1372 | 0.019 | 0.521 |
| BN_PM_Hipp_cHipp | 133.19 | 4.20 | 0.0950 | 0.042 | 1.000 |
| BN_PM_MFG_A9_46d | 130.76 | 3.17 | -0.0819 | 0.077 | 1.000 |
| BN_PM_IPL_A39rv | 112.62 | 2.77 | -0.0712 | 0.099 | 1.000 |
| BN_PM_LOcC_OPC | 133.69 | 2.72 | 0.0874 | 0.101 | 1.000 |
| BN_PM_PhG_TL | 130.49 | 2.43 | 0.0871 | 0.121 | 1.000 |
| BN_PM_MFG_A6vl | 128.77 | 2.26 | -0.0625 | 0.135 | 1.000 |
| BN_PM_MVOcC_cCunG | 129.00 | 2.09 | 0.0760 | 0.151 | 1.000 |
| BN_PM_Hipp_rHipp | 135.93 | 1.82 | 0.0851 | 0.179 | 1.000 |
| BN_PM_PCL_A1_2_3ll | 125.28 | 1.43 | 0.0418 | 0.233 | 1.000 |
| BN_PM_IPL_A39c | 129.61 | 1.33 | -0.0469 | 0.250 | 1.000 |

| | | | | | |
|---|---|---|---|---|---|
| BN_PM_ITG_A37vl | 130.56 | 1.19 | 0.0635 | 0.278 | 1.000 |
| BN_PM_LOcC_V5_MT | 132.13 | 1.18 | 0.0500 | 0.280 | 1.000 |
| BN_PM_CG_A32sg | 127.64 | 1.05 | -0.0489 | 0.306 | 1.000 |
| BN_PM_OrG_A14m | 131.88 | 1.00 | -0.0447 | 0.320 | 1.000 |
| BN_PM_PCun_A31 | 120.65 | 0.91 | -0.0384 | 0.342 | 1.000 |
| BN_PM_PCun_dmPOS | 133.23 | 0.77 | -0.0469 | 0.382 | 1.000 |
| BN_PM_MVOcC_rCunG | 132.12 | 0.73 | 0.0364 | 0.393 | 1.000 |
| BN_PM_PCun_A7m | 134.58 | 0.57 | -0.0315 | 0.450 | 1.000 |
| BN_PM_SPL_A7r | 129.51 | 0.49 | 0.0256 | 0.485 | 1.000 |
| BN_PM_CG_A23v | 131.20 | 0.48 | -0.0359 | 0.488 | 1.000 |
| BN_PM_CG_A23d | 126.57 | 0.33 | -0.0247 | 0.565 | 1.000 |
| BN_PM_LOcC_lsOccG | 129.19 | 0.30 | -0.0230 | 0.588 | 1.000 |
| BN_PM_PCun_A5m | 132.03 | 0.07 | -0.0077 | 0.794 | 1.000 |
| BN_PM_OrG_A11m | 129.01 | 0.04 | 0.0092 | 0.844 | 1.000 |
| BN_PM_MVOcC_vmPOS | 132.75 | 0.03 | 0.0072 | 0.865 | 1.000 |
| BN_PM_PCL_A4ll | 200.00 | 0.00 | 0.0015 | 0.960 | 1.000 |
| BN_PM_LOcC_msOccG | 127.25 | 0.00 | 0.0006 | 0.989 | 1.000 |

**Supplementary Table 9. Functional changes related to glucose metabolism in each region of interest (ROI) within the posterior-medial (PM) network.** Statistical summary of glucose metabolism term from linear mixed models adjusted for age, sex, education and time as F-test result. Trend was calculated as the trend of the estimated marginal means for functional changes in relation to glucose metabolism. Bolded *p*-values survived the Holm-Bonferroni correction ($p < 0.05$). Ddf: approximate denominator degrees of freedom.

| | ddf | *F* | trend | *P* | adjusted *P* |
|---|---|---|---|---|---|
| BN_PM_MVOcC_rLinG | 126.98 | 17.31 | -0.1070 | 0.000 | **0.002** |
| BN_PM_FuG_A37mv | 130.95 | 13.31 | -0.0914 | 0.000 | **0.015** |
| BN_PM_FuG_A37lv | 124.01 | 12.24 | -0.0752 | 0.001 | **0.025** |
| BN_PM_Tha_Otha | 117.73 | 11.58 | -0.0776 | 0.001 | **0.035** |
| BN_PM_LOcC_mOccG | 119.74 | 11.38 | -0.0822 | 0.001 | **0.037** |
| BN_PM_LOcC_iOccG | 124.90 | 10.38 | -0.0834 | 0.002 | 0.058 |
| BN_PM_LOcC_V5_MT | 127.36 | 9.39 | -0.0707 | 0.003 | 0.093 |
| BN_PM_PhG_A35_36c | 135.92 | 9.25 | -0.0854 | 0.003 | 0.096 |
| BN_PM_MVOcC_cLinG | 119.94 | 9.13 | -0.0751 | 0.003 | 0.101 |
| BN_PM_FuG_A20rv | 130.70 | 9.06 | -0.0867 | 0.003 | 0.101 |
| BN_PM_PhG_TL | 128.11 | 8.29 | -0.0793 | 0.005 | 0.145 |
| BN_PM_PhG_TH | 134.74 | 8.24 | -0.0841 | 0.005 | 0.145 |
| BN_PM_SFG_A8dl | 122.41 | 7.72 | 0.0539 | 0.006 | 0.183 |
| BN_PM_MFG_A8vl | 121.70 | 5.73 | 0.0512 | 0.018 | 0.510 |
| BN_PM_Hipp_cHipp | 130.69 | 4.57 | -0.0505 | 0.034 | 0.928 |

| | | | | | |
|---|---|---|---|---|---|
| BN_PM_LOcC_OPC | 126.21 | 3.93 | -0.0529 | 0.050 | 1.000 |
| BN_PM_IPL_A39rd | 125.45 | 3.45 | 0.0532 | 0.066 | 1.000 |
| BN_PM_PCL_A1_2_3ll | 123.12 | 3.17 | -0.0318 | 0.078 | 1.000 |
| BN_PM_SPL_A7c | 137.20 | 2.06 | 0.0324 | 0.153 | 1.000 |
| BN_PM_MFG_A6vl | 125.56 | 1.69 | 0.0282 | 0.196 | 1.000 |
| BN_PM_MVOcC_cCunG | 119.97 | 1.52 | -0.0324 | 0.221 | 1.000 |
| BN_PM_PCL_A4ll | 212.00 | 1.42 | -0.0180 | 0.235 | 1.000 |
| BN_PM_CG_A23d | 127.75 | 1.42 | 0.0264 | 0.236 | 1.000 |
| BN_PM_MVOcC_rCunG | 122.39 | 1.25 | -0.0234 | 0.266 | 1.000 |
| BN_PM_Hipp_rHipp | 135.04 | 1.23 | -0.0356 | 0.269 | 1.000 |
| BN_PM_MFG_A9_46d | 128.51 | 1.02 | 0.0242 | 0.314 | 1.000 |
| BN_PM_PCun_A7m | 130.08 | 0.75 | 0.0187 | 0.387 | 1.000 |
| BN_PM_ITG_A37vl | 124.45 | 0.65 | -0.0229 | 0.423 | 1.000 |
| BN_PM_LOcC_msOccG | 128.60 | 0.56 | -0.0185 | 0.457 | 1.000 |
| BN_PM_OrG_A14m | 133.81 | 0.25 | 0.0119 | 0.619 | 1.000 |
| BN_PM_CG_A32sg | 128.43 | 0.23 | 0.0122 | 0.635 | 1.000 |
| BN_PM_PCun_A5m | 128.30 | 0.14 | -0.0057 | 0.708 | 1.000 |
| BN_PM_CG_A23v | 127.65 | 0.13 | -0.0091 | 0.724 | 1.000 |
| BN_PM_SPL_A7r | 131.11 | 0.11 | 0.0066 | 0.736 | 1.000 |
| BN_PM_LOcC_lsOccG | 134.20 | 0.11 | -0.0072 | 0.742 | 1.000 |
| BN_PM_OrG_A11m | 135.29 | 0.08 | -0.0071 | 0.781 | 1.000 |
| BN_PM_IPL_A39rv | 116.68 | 0.07 | 0.0061 | 0.786 | 1.000 |
| BN_PM_IPL_A39c | 128.09 | 0.07 | -0.0055 | 0.794 | 1.000 |
| BN_PM_MVOcC_vmPOS | 125.89 | 0.03 | -0.0038 | 0.859 | 1.000 |
| BN_PM_PCun_A31 | 122.91 | 0.03 | 0.0035 | 0.870 | 1.000 |
| BN_PM_PCun_dmPOS | 132.11 | 0.00 | 0.0013 | 0.961 | 1.000 |

**Supplementary Table 10. Functional changes related to hippocampal volume in each region of interest (ROI) within the posterior-medial (PM) network.** Statistical summary of hippocampal volume term from linear mixed models adjusted for age, sex, education and time as F-test result. Trend was calculated as the trend of the estimated marginal means for functional changes in relation to hippocampal volume. Bolded *p*-values survived the Holm-Bonferroni correction ($p < 0.05$). Ddf: approximate denominator degrees of freedom.

| | ddf | *F* | trend | *P* | adjusted *P* |
|---|---|---|---|---|---|
| BN_PM_LOcC_iOccG | 115.80 | 7.73 | -0.0707 | 0.006 | 0.260 |
| BN_PM_PhG_TL | 113.94 | 5.77 | -0.0669 | 0.018 | 0.698 |
| BN_PM_MVOcC_rLinG | 116.32 | 5.98 | -0.0665 | 0.016 | 0.640 |
| BN_PM_LOcC_mOccG | 114.41 | 4.66 | -0.0554 | 0.033 | 1.000 |
| BN_PM_MVOcC_cLinG | 110.68 | 4.77 | -0.0534 | 0.031 | 1.000 |
| BN_PM_FuG_A37mv | 121.28 | 3.97 | -0.0524 | 0.048 | 1.000 |

| | | | | | |
|---|---|---|---|---|---|
| BN_PM_Tha_Otha | 114.40 | 4.54 | -0.0505 | 0.035 | 1.000 |
| BN_PM_PhG_A35_36c | 119.70 | 2.96 | -0.0486 | 0.088 | 1.000 |
| BN_PM_PhG_TH | 119.38 | 1.45 | -0.0368 | 0.230 | 1.000 |
| BN_PM_LOcC_OPC | 114.82 | 1.97 | -0.0368 | 0.164 | 1.000 |
| BN_PM_Hipp_rHipp | 119.40 | 1.12 | -0.0336 | 0.292 | 1.000 |
| BN_PM_Hipp_cHipp | 115.70 | 2.01 | -0.0333 | 0.159 | 1.000 |
| BN_PM_FuG_A20rv | 118.69 | 1.26 | -0.0330 | 0.264 | 1.000 |
| BN_PM_LOcC_V5_MT | 118.58 | 1.67 | -0.0305 | 0.199 | 1.000 |
| BN_PM_MVOcC_cCunG | 110.29 | 1.35 | -0.0294 | 0.248 | 1.000 |
| BN_PM_PCL_A1_2_3ll | 113.48 | 2.59 | -0.0288 | 0.110 | 1.000 |
| BN_PM_FuG_A37lv | 114.67 | 0.86 | -0.0206 | 0.357 | 1.000 |
| BN_PM_LOcC_msOccG | 113.27 | 0.30 | -0.0139 | 0.584 | 1.000 |
| BN_PM_PCL_A4ll | 122.22 | 0.46 | -0.0098 | 0.501 | 1.000 |
| BN_PM_ITG_A37vl | 106.13 | 0.08 | -0.0081 | 0.778 | 1.000 |
| BN_PM_LOcC_lsOccG | 117.62 | 0.03 | -0.0036 | 0.868 | 1.000 |
| BN_PM_OrG_A11m | 119.69 | 0.01 | -0.0021 | 0.933 | 1.000 |
| BN_PM_SPL_A7r | 116.21 | 0.00 | 0.0014 | 0.945 | 1.000 |
| BN_PM_MVOcC_rCunG | 115.66 | 0.01 | 0.0022 | 0.918 | 1.000 |
| BN_PM_OrG_A14m | 122.05 | 0.05 | 0.0051 | 0.830 | 1.000 |
| BN_PM_PCun_A5m | 118.06 | 0.13 | 0.0055 | 0.715 | 1.000 |
| BN_PM_CG_A32sg | 116.13 | 0.12 | 0.0086 | 0.734 | 1.000 |
| BN_PM_MFG_A6vl | 111.50 | 0.25 | 0.0111 | 0.620 | 1.000 |
| BN_PM_CG_A23d | 113.10 | 0.49 | 0.0156 | 0.486 | 1.000 |
| BN_PM_MVOcC_vmPOS | 122.27 | 0.53 | 0.0158 | 0.468 | 1.000 |
| BN_PM_PCun_dmPOS | 118.69 | 0.41 | 0.0175 | 0.522 | 1.000 |
| BN_PM_IPL_A39c | 116.49 | 0.74 | 0.0184 | 0.391 | 1.000 |
| BN_PM_MFG_A8vl | 116.18 | 1.02 | 0.0223 | 0.315 | 1.000 |
| BN_PM_PCun_A31 | 111.04 | 1.44 | 0.0251 | 0.232 | 1.000 |
| BN_PM_CG_A23v | 120.35 | 1.12 | 0.0273 | 0.293 | 1.000 |
| BN_PM_MFG_A9_46d | 119.88 | 1.43 | 0.0294 | 0.234 | 1.000 |
| BN_PM_IPL_A39rv | 107.48 | 2.14 | 0.0325 | 0.146 | 1.000 |
| BN_PM_IPL_A39rd | 115.51 | 1.36 | 0.0329 | 0.245 | 1.000 |
| BN_PM_PCun_A7m | 117.58 | 3.11 | 0.0375 | 0.080 | 1.000 |
| BN_PM_SFG_A8dl | 116.79 | 3.85 | 0.0391 | 0.052 | 1.000 |
| BN_PM_SPL_A7c | 121.76 | 3.96 | 0.0448 | 0.049 | 1.000 |

**Supplementary Table 11. Functional changes related to Mini-Mental Scale Examination score (MMSE) in each region of interest (ROI) within the posterior-medial (PM) network.** Statistical summary of MMSE term from linear mixed models adjusted for age, sex, education and time as F-test result. Trend was calculated as the trend of the estimated marginal means for functional changes in relation to MMSE. Bolded *p*-values survived the Holm-Bonferroni correction ($p < 0.05$). Ddf: approximate denominator degrees of freedom.

| | ddf | *F* | trend | *P* | adjusted *P* |
|---|---|---|---|---|---|
| BN_PM_FuG_A37mv | 163.71 | 18.94 | -0.0071 | 0.000 | **0.001** |
| BN_PM_MVOcC_rLinG | 158.80 | 14.06 | -0.0064 | 0.000 | **0.009** |
| BN_PM_LOcC_iOccG | 158.41 | 14.53 | -0.0062 | 0.000 | **0.008** |
| BN_PM_PhG_A35_36c | 162.54 | 11.31 | -0.0059 | 0.001 | **0.036** |
| BN_PM_FuG_A20rv | 162.90 | 9.65 | -0.0057 | 0.002 | 0.076 |
| BN_PM_FuG_A37lv | 158.19 | 16.46 | -0.0056 | 0.000 | **0.003** |
| BN_PM_Tha_Otha | 160.87 | 11.31 | -0.0052 | 0.001 | **0.036** |
| BN_PM_MVOcC_cLinG | 155.37 | 9.76 | -0.0050 | 0.002 | 0.074 |
| BN_PM_PhG_TH | 165.32 | 5.98 | -0.0046 | 0.016 | 0.451 |
| BN_PM_LOcC_mOccG | 157.07 | 7.97 | -0.0045 | 0.005 | 0.172 |
| BN_PM_LOcC_OPC | 158.31 | 6.81 | -0.0044 | 0.010 | 0.297 |
| BN_PM_PhG_TL | 159.37 | 5.37 | -0.0041 | 0.022 | 0.589 |
| BN_PM_Hipp_cHipp | 160.63 | 7.04 | -0.0040 | 0.009 | 0.272 |
| BN_PM_Hipp_rHipp | 164.75 | 3.29 | -0.0036 | 0.071 | 1.000 |
| BN_PM_LOcC_V5_MT | 161.16 | 3.97 | -0.0030 | 0.048 | 1.000 |
| BN_PM_MVOcC_cCunG | 154.04 | 2.78 | -0.0028 | 0.098 | 1.000 |
| BN_PM_ITG_A37vl | 153.48 | 2.02 | -0.0026 | 0.157 | 1.000 |
| BN_PM_PCL_A1_2_3ll | 157.06 | 4.01 | -0.0024 | 0.047 | 1.000 |
| BN_PM_OrG_A11m | 166.93 | 0.42 | -0.0010 | 0.519 | 1.000 |
| BN_PM_MVOcC_rCunG | 159.34 | 0.51 | -0.0010 | 0.474 | 1.000 |
| BN_PM_PCL_A4ll | 167.72 | 0.59 | -0.0008 | 0.444 | 1.000 |
| BN_PM_CG_A23v | 163.45 | 0.03 | -0.0003 | 0.867 | 1.000 |
| BN_PM_PCun_A31 | 158.37 | 0.01 | -0.0001 | 0.941 | 1.000 |
| BN_PM_CG_A32sg | 163.39 | 0.02 | 0.0002 | 0.888 | 1.000 |
| BN_PM_LOcC_lsOccG | 172.49 | 0.03 | 0.0002 | 0.856 | 1.000 |
| BN_PM_LOcC_msOccG | 165.95 | 0.03 | 0.0003 | 0.864 | 1.000 |
| BN_PM_OrG_A14m | 169.94 | 0.05 | 0.0003 | 0.817 | 1.000 |
| BN_PM_PCun_A5m | 161.35 | 0.20 | 0.0004 | 0.657 | 1.000 |
| BN_PM_IPL_A39c | 161.69 | 0.19 | 0.0006 | 0.661 | 1.000 |
| BN_PM_MVOcC_vmPOS | 165.52 | 0.26 | 0.0007 | 0.612 | 1.000 |
| BN_PM_SPL_A7r | 160.57 | 0.57 | 0.0009 | 0.451 | 1.000 |
| BN_PM_CG_A23d | 158.27 | 0.49 | 0.0010 | 0.486 | 1.000 |
| BN_PM_IPL_A39rv | 153.63 | 0.96 | 0.0014 | 0.330 | 1.000 |
| BN_PM_PCun_dmPOS | 163.50 | 1.04 | 0.0018 | 0.308 | 1.000 |
| BN_PM_MFG_A6vl | 158.11 | 1.93 | 0.0019 | 0.166 | 1.000 |
| BN_PM_MFG_A9_46d | 164.11 | 1.55 | 0.0020 | 0.215 | 1.000 |

| | | | | | |
|---|---|---|---|---|---|
| BN_PM_PCun_A7m | 160.56 | 2.84 | 0.0023 | 0.094 | 1.000 |
| BN_PM_MFG_A8vl | 160.31 | 4.04 | 0.0029 | 0.046 | 1.000 |
| BN_PM_SPL_A7c | 166.27 | 5.50 | 0.0034 | 0.020 | 0.565 |
| BN_PM_IPL_A39rd | 160.78 | 3.93 | 0.0036 | 0.049 | 1.000 |
| BN_PM_SFG_A8dl | 160.47 | 9.20 | 0.0039 | 0.003 | 0.093 |

__Supplementary Table 12.__ **Functional changes related to Mattis Dementia Rating Scale score (MDRS) in each region of interest (ROI) within the posterior-medial (PM) network.** Statistical summary of MDRS term from linear mixed models adjusted for age, sex, education and time as F-test result. Trend was calculated as the trend of the estimated marginal means for functional changes in relation to MDRS. Bolded *p*-values survived the Holm-Bonferroni correction ($p < 0.05$). Ddf: approximate denominator degrees of freedom.

| | ddf | *F* | trend | *P* | adjusted *P* |
|---|---|---|---|---|---|
| BN_PM_PhG_A35_36c | 158.56 | 9.62 | -0.0021 | 0.002 | 0.089 |
| BN_PM_FuG_A37mv | 166.43 | 10.36 | -0.0020 | 0.002 | 0.064 |
| BN_PM_MVOcC_rLinG | 162.86 | 9.17 | -0.0020 | 0.003 | 0.100 |
| BN_PM_LOcC_iOccG | 157.78 | 9.49 | -0.0019 | 0.002 | 0.090 |
| BN_PM_Tha_Otha | 155.58 | 8.91 | -0.0018 | 0.003 | 0.112 |
| BN_PM_PhG_TH | 164.05 | 5.96 | -0.0017 | 0.016 | 0.504 |
| BN_PM_FuG_A37lv | 162.23 | 10.26 | -0.0017 | 0.002 | 0.066 |
| BN_PM_MVOcC_cLinG | 155.78 | 7.64 | -0.0017 | 0.006 | 0.211 |
| BN_PM_Hipp_cHipp | 158.24 | 5.81 | -0.0014 | 0.017 | 0.530 |
| BN_PM_LOcC_OPC | 156.52 | 4.49 | -0.0014 | 0.036 | 1.000 |
| BN_PM_FuG_A20rv | 163.02 | 3.63 | -0.0014 | 0.058 | 1.000 |
| BN_PM_MVOcC_cCunG | 152.40 | 3.22 | -0.0011 | 0.075 | 1.000 |
| BN_PM_LOcC_mOccG | 158.91 | 3.12 | -0.0011 | 0.079 | 1.000 |
| BN_PM_PhG_TL | 160.22 | 2.20 | -0.0010 | 0.140 | 1.000 |
| BN_PM_ITG_A37vl | 157.01 | 1.24 | -0.0008 | 0.267 | 1.000 |
| BN_PM_Hipp_rHipp | 164.89 | 0.97 | -0.0008 | 0.325 | 1.000 |
| BN_PM_LOcC_V5_MT | 160.97 | 1.06 | -0.0006 | 0.304 | 1.000 |
| BN_PM_PCL_A1_2_3ll | 162.93 | 1.54 | -0.0005 | 0.216 | 1.000 |
| BN_PM_OrG_A11m | 167.88 | 0.26 | -0.0003 | 0.611 | 1.000 |
| BN_PM_MVOcC_rCunG | 160.00 | 0.33 | -0.0003 | 0.567 | 1.000 |
| BN_PM_PCL_A4ll | 173.39 | 0.00 | 0.0000 | 0.989 | 1.000 |
| BN_PM_CG_A23v | 165.72 | 0.00 | 0.0000 | 0.963 | 1.000 |
| BN_PM_CG_A32sg | 164.88 | 0.04 | 0.0001 | 0.845 | 1.000 |
| BN_PM_PCun_A31 | 162.19 | 0.22 | 0.0002 | 0.643 | 1.000 |
| BN_PM_LOcC_lsOccG | 170.57 | 0.37 | 0.0003 | 0.543 | 1.000 |
| BN_PM_PCun_A5m | 167.05 | 1.06 | 0.0004 | 0.306 | 1.000 |
| BN_PM_LOcC_msOccG | 163.58 | 0.52 | 0.0004 | 0.474 | 1.000 |

| | | | | | |
|---|---|---|---|---|---|
| BN_PM_OrG_A14m | 170.72 | 0.54 | 0.0004 | 0.465 | 1.000 |
| BN_PM_CG_A23d | 163.18 | 0.88 | 0.0005 | 0.348 | 1.000 |
| BN_PM_IPL_A39c | 161.79 | 1.07 | 0.0005 | 0.303 | 1.000 |
| BN_PM_MVOcC_vmPOS | 161.25 | 1.06 | 0.0006 | 0.304 | 1.000 |
| BN_PM_SPL_A7r | 162.31 | 1.53 | 0.0006 | 0.218 | 1.000 |
| BN_PM_MFG_A6vl | 162.50 | 2.36 | 0.0008 | 0.126 | 1.000 |
| BN_PM_IPL_A39rv | 158.52 | 2.36 | 0.0008 | 0.126 | 1.000 |
| BN_PM_MFG_A9_46d | 168.23 | 2.17 | 0.0009 | 0.142 | 1.000 |
| BN_PM_PCun_A7m | 164.66 | 2.83 | 0.0009 | 0.095 | 1.000 |
| BN_PM_PCun_dmPOS | 164.64 | 2.08 | 0.0009 | 0.151 | 1.000 |
| BN_PM_MFG_A8vl | 164.78 | 5.33 | 0.0012 | 0.022 | 0.666 |
| BN_PM_IPL_A39rd | 163.89 | 3.34 | 0.0013 | 0.070 | 1.000 |
| BN_PM_SFG_A8dl | 165.21 | 9.59 | 0.0015 | 0.002 | 0.089 |
| BN_PM_SPL_A7c | 168.36 | 9.32 | 0.0016 | 0.003 | 0.095 |

**Supplementary Table 13. Statistical summary of replication analyses across the whole sample, covering the entire adult lifespan and the entire cognitive continuum from normal to dementia, using connectivity values extracted from young adults' masks**. The *group* term was derived from generalized additive models on baseline data. Visual inspection of the smooth term indicates similar trajectories to those illustrated in Fig. 6. The *time* x *group* interaction term was derived from mixed models using longitudinal data. Visual inspection of post-hoc predictions indicates similar effects to those depicted in Fig. 6.

| *Group* | | | |
|---|---|---|---|
| | edf | *F* | *P* |
| AT | 1.56 | 13 | **<0.001** |
| PM | 2.27 | 9.28 | **<0.001** |

| *Time * Group* | | | |
|---|---|---|---|
| | ddf | *F* | *P* |
| AT | 310.13 | 4.64 | **0.032** |
| PM | 325.87 | 1.03 | 0.312 |

**Supplementary Table 14. Statistical summary of complementary analyses including both networks in the same model.** All outputs were consistent with statistics reported in Results.

| Interaction with network | | | | Post-hoc | | | | | |
|---|---|---|---|---|---|---|---|---|---|
| | | | | AT | | | PM | | |
| | ddf | *F* | *P* | trend | t | *P* | trend | t | *P* |
| Age | 624.8 | 42.6 | **<0.001** | $2.99e^{-4}$ | 2.3 | **0.023** | $-7.08\ e^{-4}$ | -5.4 | **<0.001** |
| Amyloid uptake | 304.8 | 8.6 | **0.004** | $1.62e^{-1}$ | 4.8 | **<0.001** | $3.86e^{-2}$ | 1.1 | 0.255 |

| | | | | | | | | | |
|---|---|---|---|---|---|---|---|---|---|
| Glucose metabolism | 328.7 | 15.3 | **<0.001** | $-1.07e^{-1}$ | -6.5 | **<0.001** | $-2.54e^{-2}$ | -1.5 | 0.127 |
| Hippocampal volume | 356.6 | 25.9 | **<0.001** | $-8.64e^{-2}$ | -5.1 | **<0.001** | $1.21e^{-2}$ | 0.7 | 0.472 |
| MMSE | 359.5 | 5.3 | **0.022** | $-4.95e^{-3}$ | -4.4 | **<0.001** | $-1.79e^{-3}$ | -1.6 | 0.109 |
| MDRS | 334.5 | 8.0 | **0.005** | $-1.87e^{-3}$ | -4.5 | **<0.001** | $-4.11e^{-4}$ | -0.98 | 0.329 |
| MCI-to-AD time | 69 | 13.0 | **<0.001** | $-1.56e^{-2}$ | -3.3 | **0.002** | $7.11e^{-3}$ | 1.5 | 0.141 |
